# Identification of circulating lipidomic biomarkers of malnutrition risk among oncology patients in the Total Cancer Care (TCC) Study: a cross-sectional analysis

**DOI:** 10.64898/2026.01.09.26343797

**Authors:** Rachel Hoobler, J Alan Maschek, Bai Luo, E. Angela Murphy, Jason L. Kubinak, Paul A. Stewart, James E Cox, Amandine Chaix, Kary Woodruff, Alejandro Sanchez, Adriana M Coletta, Fred K. Tabung, Sumati Gupta, Sheetal Hardikar, Howard Colman, Mary C Playdon

**Affiliations:** Department of Nutrition & Integrative Physiology, University of Utah, Salt Lake City, UT, USA; Huntsman Cancer Institute, University of Utah, Salt Lake City, UT, USA; Diabetes and Metabolism Research Center, University of Utah, Salt Lake City, UT, USA; Metabolomics Core Research Facility, University of Utah, Salt Lake City, UT, USA; Drug Discovery Core Facility, University of Utah, Salt Lake City, UT, USA; Department of Pathology, Microbiology and Immunology, School of Medicine, University of South Carolina, Columbia, SC, USA; Department of Biochemistry, University of Utah, Salt Lake City, UT, USA; Division of Urology, Department of Surgery, Huntsman Cancer Institute, University of Utah, Salt Lake City, UT, USA; Department of Health and Kinesiology, University of Utah, Salt Lake City, UT, USA; The Ohio State University Comprehensive Cancer Center, Columbus, OH, USA; Division of Medical Oncology, Department of Internal Medicine, College of Medicine, The Ohio State University, Columbus, OH, USA; Department of Internal Medicine, Huntsman Cancer Institute, University of Utah, Salt Lake City, UT, USA; George E. Wahlen Department of Veterans Affairs Medical Center, Salt Lake City, UT, USA; Department of Population Health Sciences, University of Utah, Salt Lake City, UT, USA; Department of Neurosurgery, School of Medicine, University of Utah, Salt Lake City, UT, USA

**Keywords:** Lipidomics, Malnutrition, Biomarkers, Oncology, Cancer

## Abstract

**Background:** Early identification of malnutrition is critical for improving clinical outcomes in oncology patients. However, there are no established biomarkers for malnutrition screening.

**Objective:** This study aimed to identify circulating lipid species associated with malnutrition risk among oncology patients through lipidomic analysis.

**Methods:** A cross-sectional study was conducted using plasma samples from oncology patients classified as at risk (n = 90) or not at risk (n = 90) for malnutrition using the Malnutrition Screening Tool (MST) (MST score = 0 versus ≥2). All participants had head and neck, lung, or gastrointestinal cancer. Targeted lipidomics were conducted using LC-MS. Elastic net regression adjusted for confounding variables identified lipids associated with malnutrition risk. A weighted Lipid Malnutrition Risk Score was derived and evaluated using Receiver Operating Characteristic Area Under the Curve (ROC-AUC). Conditional multivariable logistic regression assessed the association of the lipid score with malnutrition risk. Lipid enrichment analysis was performed using Lipid Ontology (LION) enrichment framework.

**Results:** Elastic net regression identified 12 lipids species that were inversely associated with malnutrition risk: cholesterol ester 20:0, ceramide 18:2;O2/26:0, lysophosphatidylcholine 26:0/0:0, lysophosphatidylinositol 18:2/0:0, phosphatidylcholine 34:5, phosphatidylcholine 40:8, phosphatidylethanolamine P-18:0/20:3, phosphatidylethanolamine P-18:1/18:2, phosphatidylethanolamine P-18:1/20:4, sulfated hexosylceramide 18:1;O2/16:0, sphingomyelin 18:2;O2/23:0, and triglyceride (O-50:1). One lipid, dihexosylceramide 18:1;O2/20:0, was positively associated with malnutrition risk. The weighted Lipid Malnutrition Risk Score was associated with increased risk for malnutrition risk (OR = 3.57, 95% CI 1.97-6.47, p < 0.001). Addition of the lipids score to established malnutrition risk factors improved model predictive performance, increasing the ROC-AUC from 0.78 (95% CI 0.71-0.84) to 0.90 (95% CI 0.86-0.94). LION enrichment analysis indicated downregulation of membrane structure and signaling lipids and upregulation of storage lipids.

**Conclusion:** This study highlights the potential of lipidomics to identify biomarkers of malnutrition risk among oncology patients. Large, prospective studies are warranted to validate and expand upon these findings.

## Introduction

Malnutrition is prevalent among oncology patients, with estimates suggesting that up to 80% of patients with cancer experience malnutrition during the course of treatment (1–7). This is concerning as untreated malnutrition before or during treatment is associated with increased risk for post-operative complications, treatment toxicity, reduced quality of life, higher likelihood of cancer recurrence, and worse survival outcomes (8–16). Additionally, cancer-related malnutrition imposes a substantial financial burden, as undernourished patients often incur greater healthcare costs, largely due to prolonged hospital stays (6, 15, 17–20).

Malnutrition screening refers to quick and simple processes used to identify patients at risk for malnutrition who may benefit from a comprehensive nutritional assessment by a Registered Dietitian (**RD**). Early malnutrition screening, followed by timely referral to an RD for patients identified as at risk, is recommended to prevent and mitigate the consequences of malnutrition (21). Nutritional interventions provided by an RD have been shown to improve calorie and protein intake, quality of life, and anthropometric measures among oncology patients, as well as, reduced length of hospital stays and costs (22–24).

Several validated malnutrition screening tools exist for both inpatient and outpatient use (25). However, these screening tools rely primarily on patient-reported information, such as changes in appetite or food intake and unintentional weight loss (26–29). These criteria are subjective and prone to misclassification of malnutrition risk status. Multiple screening tools also incorporate body mass index (**BMI**), which may fail to detect malnutrition risk in overweight and obese patients (26, 27, 30). To date, there are no valid, sensitive, and specific biomarkers of malnutrition or malnutrition risk recommended for clinical or research use (31, 32).

Metabolomic and lipidomic technologies offer new opportunities to identify biomarkers that may enhance diagnosis and disease detection in oncology patients (33–36). Previous studies have identified potential biomarkers of malnutrition using metabolomics (37–40), but few studies have been conducted among patients with cancer, nor have they employed lipidomics (41). Lipids play a key role in cancer biology (42–44), and prior studies indicate that lipid species, such as lysophosphatidylcholines (**LPCs**), have potential as malnutrition biomarkers (41, 45–47).

Identification of valid and reliable malnutrition biomarkers could enhance the sensitivity and specificity of malnutrition screening, enabling earlier detection of poor nutritional status, timely intervention, and improved patient outcomes. This study aimed to identify a panel of lipids associated with malnutrition risk. We hypothesized that patients at risk of malnutrition per the Malnutrition Screening Tool (**MST**) (MST score ≥2) would exhibit a distinct plasma lipidomic profile compared to those without malnutrition risk (MST score = 0), and that this profile would be independently associated with malnutrition risk and improve classification of malnutrition risk status beyond established risk factors.

## Methods

A cross-sectional study was conducted using plasma samples from the Total Cancer Care (**TCC**) cohort at the Huntsman Cancer Institute (**HCI**) (48, 49) and data from electronic medical records (**EMR**). Data were abstracted from January 2021 to April 2024. Included participants had lung (International Classification of Diseases for Oncology (**ICD-O**) C33.9/C34), head and neck (ICD-O C00-C14/C30-C32), or gastrointestinal (C15-C26) cancer. These cancer types were selected based on their high prevalence of malnutrition (3, 7, 50–52). For patients with multiple cancer diagnoses, the most recent diagnosis was selected for analysis. If duplicates remained, we selected for the cancer with the higher stage, as cancer stage is associated with risk for malnutrition (7, 50, 53, 54).

Included participants from TCC had at least one plasma sample ≥150 microliters (**uL**) within ±30 days of an MST score (n = 684). For patients with more than two plasma samples within 30 days of an MST score, the sample closest to the MST score was selected. Participants were excluded if their sample had an unknown anticoagulation treatment (n = 3) or if they had limited plasma samples banked (<5 samples for controls or ≤1 sample for cases) (**Supplementary Figure 1**). After exclusions, 90 potential cases (MST score ≥2) and 490 potential controls (MST score = 0) remained. Ninety controls were then selected to match cases according to cancer type (head and neck, lung, colon, esophageal, pancreatic, rectosigmoid, small intestine, other biliary tract, or stomach), TCC ancillary study, number of freeze-thaws, and anticoagulation treatment (Ethylenediaminetetraacetic acid (**EDTA**), sodium citrate, Acid Citrate Dextrose (**ACD**) Solution A, or Cell Preparation Tube (**CPT**)) (55). This study was reviewed by the University of Utah’s Institutional Review Board (**IRB**) and was determined to be non- human subject research in accordance with federal regulations (45 CFR 46.102). Thus, IRB oversight was not required.

## Malnutrition Risk

Malnutrition risk was determined using the MST, a validated malnutrition screening tool for both inpatient and outpatient settings (28). MST was implemented in nine HCI outpatient clinics (head and neck, thoracic, gastrointestinal, sarcoma, neurology, bone marrow transplant/hematology, genitourinary, melanoma, and breast/gynecologic clinics) in January 2021. HCI outpatient clinics serve urban, rural, and frontier patients from the Mountain West, including Utah, Idaho, Montana, Nevada, and Wyoming. At the HCI outpatient centers, patients undergo malnutrition screening conducted by a medical assistant every 30 days unless they meet with an RD 15 days before or after their appointment. MST assesses changes in appetite or food intake (yes or no) and recent unintentional weight loss (no, unsure, or yes and how much). Using this information, a score between zero and five is generated. A score of ≥2 indicates malnutrition risk, while a score <2 indicates no risk (25, 28). For this analysis, patients were classified as controls if their MST score was zero, and cases if their score was ≥2, to reduce misclassification of exposure status and increase statistical power (56).

## Lipidomics Platform

The lipidomics pipeline was implemented at the University of Utah Metabolomics Core.

## Chemicals

Liquid chromatography-mass spectrometry (**LC-MS**)-grade solvents were obtained from Honeywell Burdick & Jackson (methanol; Morristown, NJ) and Fisher Scientific (1-butanol; Waltham, MA). Internal standards were purchased from Avanti Polar Lipids (Birmingham, AL), including: C16 Ceramide-d7 (d18:1-d7/16:0, 860676), C18 Ceramide-d7 (d18:1-d7/18:0, 860677), C24 Ceramide-d7 (d18:1-d7/24:0, 860678), C24:1 Ceramide-d7 (d18:1-d7/24:1(15Z), 860679), C17 Glucosyl(β)Ceramide (d18:1/17:0, 860569), C18:1 Dihydroceramide (d18:0/18:1(9Z), 860662), Sphingosine- 1-Phosphate-d7 (860659), Sphinganine-1-Phosphate-d7 (860694), Sphingosine-d7 (860657), Oleic Acid-d9 (861809), N-12:0-1-deoxysphinganine (860481), C8 Dihydroceramide (d18:0/8:0, 860626), and SPLASH Lipidomix (330707). Additional internal standards were obtained from Cayman Chemical (Ann Arbor, MI): Deoxycholic Acid-d4 (20851), Cholic Acid-d4 (20849), and d3-Palmitoylcarnitine (C16, 26569).

## Sample Preparation

Plasma samples were obtained from the Biorepository and Molecular Pathology (**BMP**) Shared Resource at the HCI and collected and stored under the TCC biospecimen collection protocol. Lipid extraction was performed using a modified version of the single-phase butanol–methanol method previously described (57, 58). Briefly, 20 µL of each sample was transferred to a 96-well plate, and lipids were extracted by adding 200 µL of a pre-mixed methanol:1-butanol solution containing internal standards. Samples were vortexed using a plate shaker, sonicated for 10 minutes at 4 °C, and centrifuged at 1,100 × g for 10 minutes at 4 °C. The resulting supernatants were transferred to new 96-well plates for LC-MS analysis. A process blank and a pooled quality control (**QC**) sample were included alongside the study samples (57).

## Lipidomics Analysis

Lipidomic analysis was performed using a liquid chromatography-tandem mass spectrometry (**LC-MS/MS**) method adapted from Huynh et al. (59), with acquisition parameters guided by an Agilent Technologies application note (60). Batch correction was performed using Systematic Error Removal using Random Forest (**SERRF**) (61), with QC samples injected at regular intervals throughout data acquisition to monitor and correct for systematic drift. SERRF was implemented in RStudio (R version 4.2.1) using a customized script from the SERRF GitHub repository, modified for the current dataset. This approach enabled correction of batch effects while preserving biological variance. A total of 701 lipids were quantified.

## Covariates

Covariates were abstracted from the EMR, including age at cancer diagnosis (continuous), biologic sex (male, female), smoking status (never, former, current), marital status (married, unmarried), BMI at first MST score (continuous), cancer stage (0/I, II, III, IV), chemotherapy (yes, no), radiation (yes, no), immunotherapy (yes, no), and surgery (yes, no). BMI data were missing for nine participants and were imputed using Predictive Mean Matching (**PMM**) (62). We were unable to adjust for race or ethnicity in this analysis due to the small number of non-White participants (i.e., Black or African American, Asian, American Indian or Alaska Native, Native Hawaiian or other Pacific Islander, and other race groups). To adjust for potential cachexia (63, 64), plasma Growth/Differentiation Factor 15 (**GDF15**) levels were measured using Human GDF15 enzyme-linked immunosorbent assay (**ELISA**) kits from R&D Systems, performed by the University of Utah Drug Discovery Core (65).

## Statistical Analysis

Participant characteristics were summarized overall and by case/control status using median and interquartile range (**IQR**) for continuous variables and counts and percentages for categorical variables. The normality of continuous variables was assessed using the Shapiro-Wilk test, and all were found to be skewed. Differences in demographic and cancer characteristics between cases and controls were evaluated using the Mann-Whitney U Test for continuous variables and the Chi Square Test of Independence for categorical variables. Fisher’s Exact Test was applied for categorical variables with cell counts less than five.

Lipid levels were normalized using an inverse normal transformation. Associations between individual lipids and malnutrition risk were assessed using conditional logistic regression. A total of 372 lipids with a Benjamini Hochberg false discovery rate (**FDR**) (66) <0.05 were retained for further analyses (67). Elastic net regression (68) was then applied to these 372 lipids associated with malnutrition risk, with age at cancer diagnosis, sex, BMI, cancer stage, and treatment with chemotherapy, radiation, surgery, or immunotherapy included as unpenalized covariates to adjust for potential confounding. Elastic net regression was run 50 times. For each iteration, the data were randomly split into training (70%) and testing (30%) sets, and the optimal regularization parameter (lambda) was determined using 10-fold cross validation. The final lipid panel included lipids selected in >90% of the iterations (69).

Three sensitivity analyses were conducted: 1) adjusting for GDF15 to distinguish lipids associated with malnutrition risk alone from those potentially driven by cachexia, given GDF15’s established link to cancer-related cachexia (63, 64); 2) adjusting for imperfectly matched matching variables (i.e., number of freeze/thaws and anticoagulation treatment) to assess for potential residual confounding; and 3) excluding patients diagnosed with cancer before 2021 when MST was implemented at HCI outpatient clinics, as although these individuals had available MST scores, they were likely long-term cancer survivors and not undergoing active treatment, which could affect their plasma lipidomic profiles.

A “Lipid Malnutrition Risk Score” was calculated by summing the elastic net selected lipids, weighted by the average beta coefficients derived from the 50 elastic net regression iterations (67, 70). Conditional logistic regression was used to assess the association between the z-score scaled lipid score and malnutrition risk, using both minimally (age, sex, and BMI) and fully (age, sex, BMI, cancer stage, smoking status, and cancer treatment) adjusted models. Covariates were selected based on their previous or hypothesized associations with malnutrition and malnutrition risk (3, 7, 50, 53, 54, 71–76). In secondary analysis, models were stratified by cancer type and cancer stage. There was inadequate statistical power to stratify by BMI.

To evaluate the predictive performance of the lipid score beyond established risk factors, we used Receiver Operating Characteristic Area Under the Curve (**ROC-AUC**) C-Statistic, Continuous Net Reclassification Improvement (**NRI**) (77), and Integrated Discrimination Improvement (**IDI**) (69, 78). Confidence intervals for IDI were calculated using standard error estimation rather than bootstrapping, due to model convergence issues. All analyses were conducted using R statistical Software (79). All tests were two-sided, and a p-value <0.05 was considered statistically significant.

Lipid enrichment analysis was preformed using Lipid Ontology (**LION**) enrichment analysis (80). Enrichment analysis identifies lipid classes and properties that differ significantly between cases and controls. Individual lipid analysis (i.e., elastic net regression) highlights lipids most predictive of malnutrition risk, while LION analysis reveals broader class-level changes that offer mechanistic insights. Raw lipid data were log2-transformed prior to enrichment analysis; this transformation was performed independently of the inverse normal transformation applied before elastic net regression. Briefly, lipids were ranked based on the results of a one-tailed Welch’s t-test, and the analysis was conducted in ranking mode to identify lipid classes and biophysical properties (e.g., membrane curvature, bilayer thickness, lateral diffusion) that were significantly enriched, as well as whether these lipid terms were up- or down- regulated in cases compared to controls. Enrichment was assessed using a one-tailed Kolmogorov-Smirnov test implemented in LION (80). All tests were two-sided and lipid terms with an FDR <0.05 were considered significantly enriched.

## Results

Patient demographic and clinical characteristics are presented in **Table 1**. Patients (n = 180) had a median (IQR) age of 65 (13) years old, over half were male (58%), and most were white (86%) and non-Hispanic or Latino (93%). The median (IQR) time from MST score to blood sample collection was zero (28.3) days. Most patients were married (66%) and either never smokers (42%) or former smokers (44%). Patients had a median (IQR) BMI of 26 (7) kg/m^2^. Nearly half of patients had head and neck cancer (49%), while 23.3% had gastrointestinal cancer and 28% had lung cancer. Thirty-two percent of patients had stage IV cancer, 23% had stage 0/I cancer, 17% had stage II cancer, and 17% had stage III cancer. Most patients underwent surgery (68%), half (51%) received chemotherapy, 39% were treated with radiation, and a small portion received immunotherapy (16%). Thirty participants (17%) were deceased at the time of data abstraction. Cases were more likely to be unmarried (p<0.01) and current or former smokers (p<0.01).

**Table 1.**
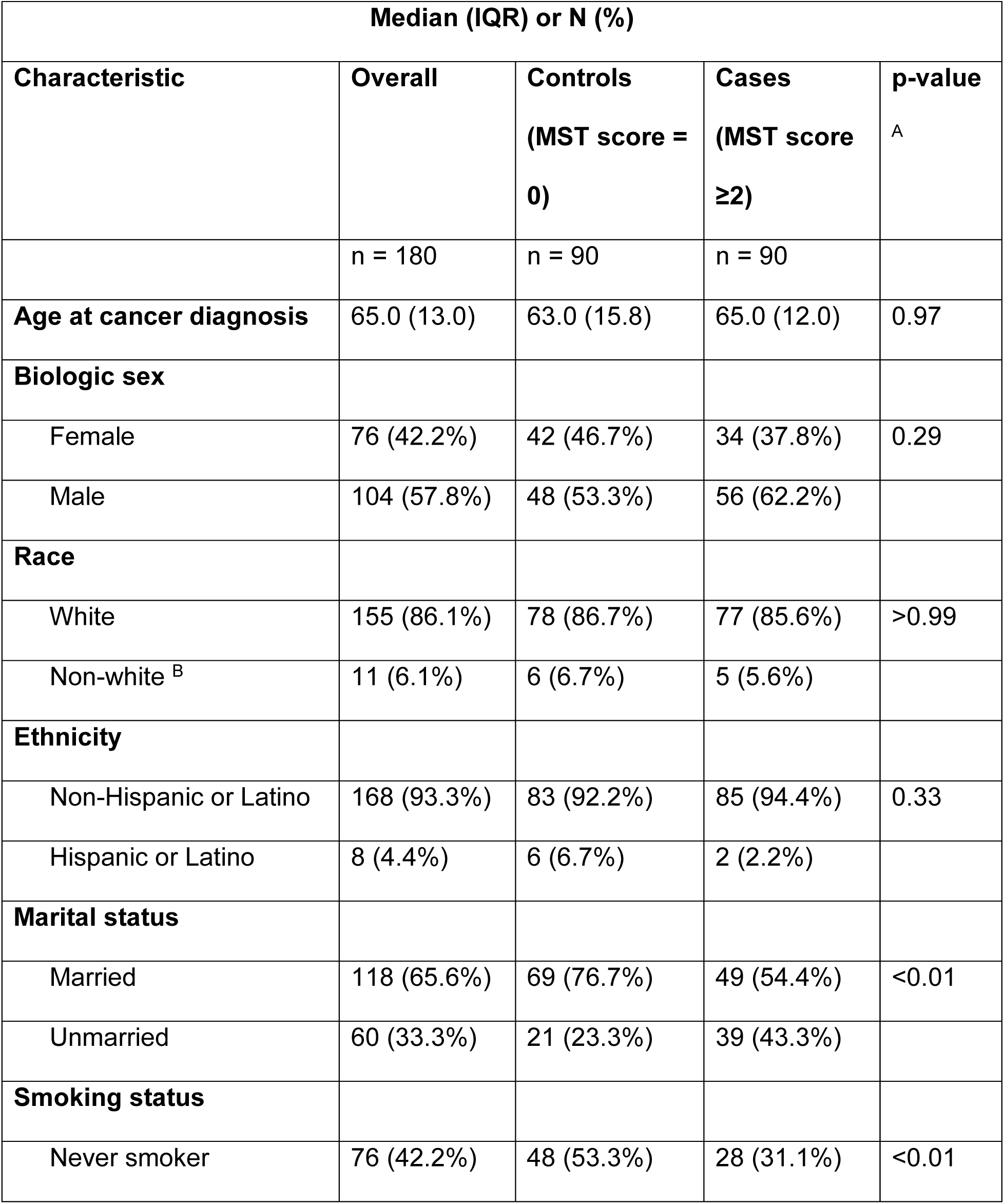
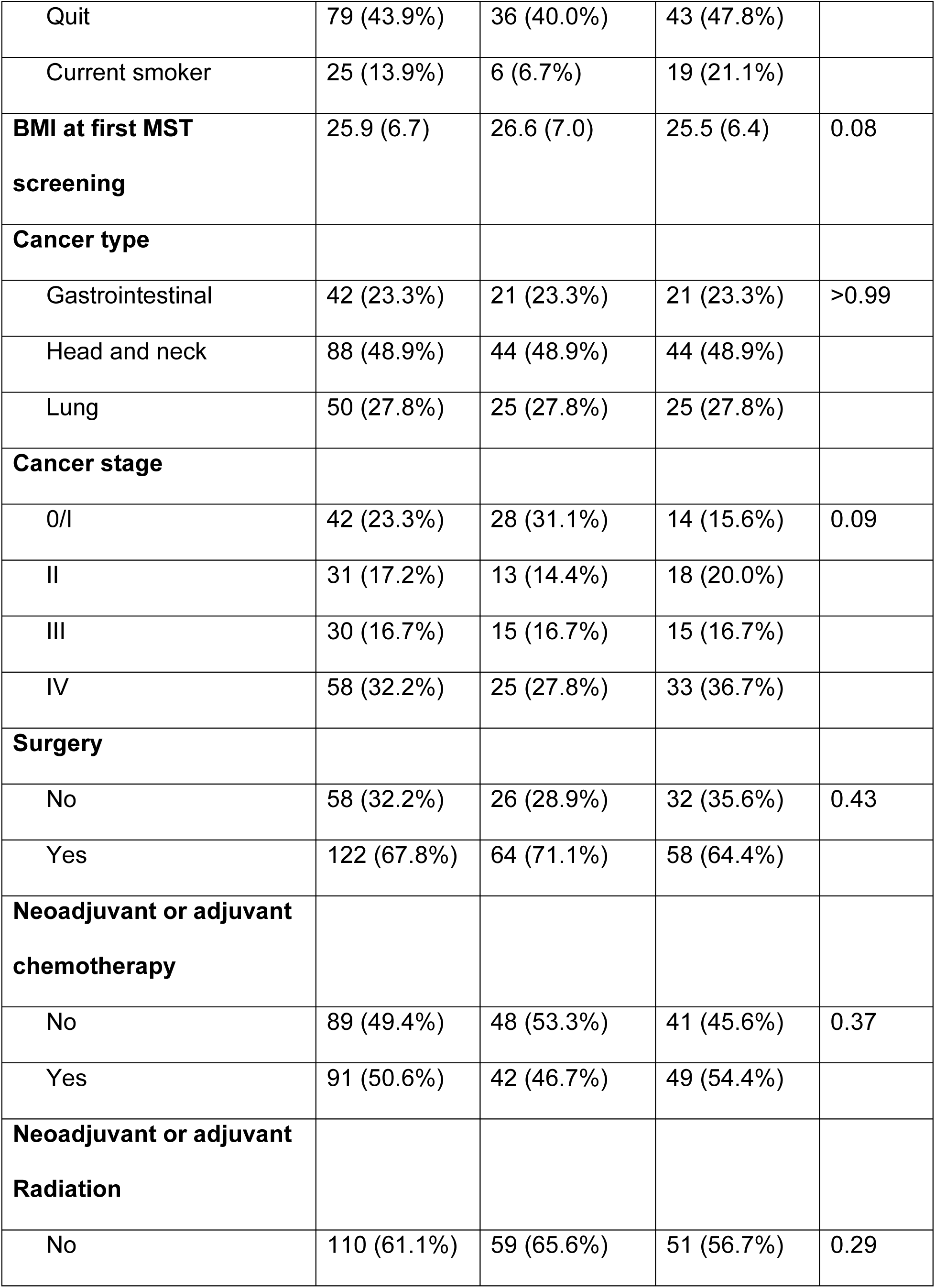
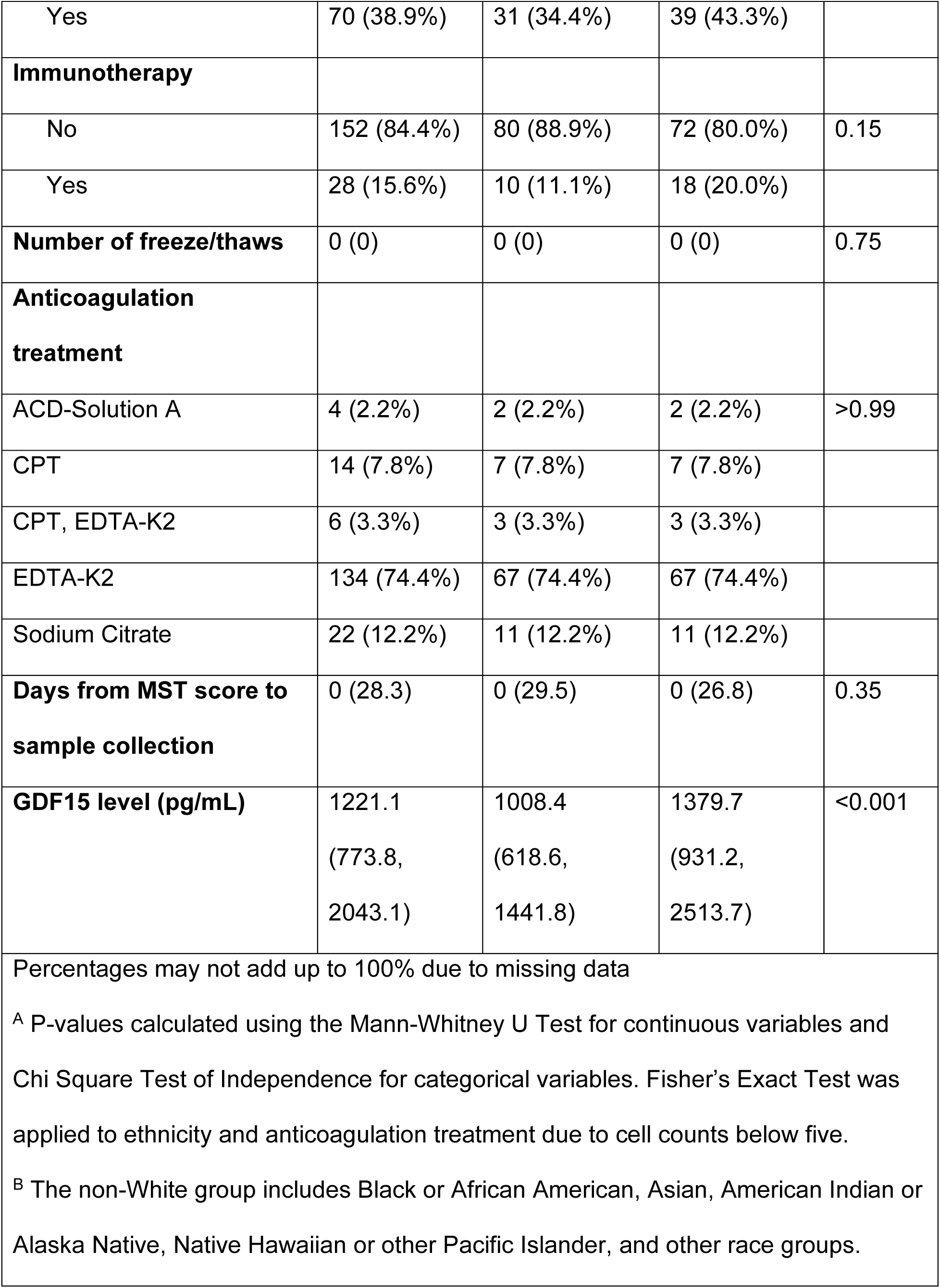
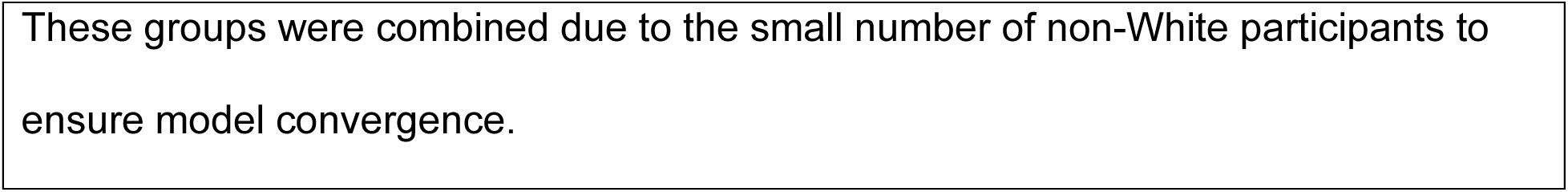
Demographic and cancer characteristics of the 180 patients included in primary analysis with head and neck, gastrointestinal, or lung cancers treated at the Huntsman Cancer Institute (HCI).

| Median (IQR) or N (%) |  |  |  |  |
| --- | --- | --- | --- | --- |
| Characteristic | Overall | Controls<br>(MST score = 0) | Cases<br>(MST score $\geq 2$) | p-value<sup>A</sup> |
|  | n = 180 | n = 90 | n = 90 |  |
| **Age at cancer diagnosis** | 65.0 (13.0) | 63.0 (15.8) | 65.0 (12.0) | 0.97 |
| **Biologic sex** |  |  |  |  |
| Female | 76 (42.2%) | 42 (46.7%) | 34 (37.8%) | 0.29 |
| Male | 104 (57.8%) | 48 (53.3%) | 56 (62.2%) |  |
| **Race** |  |  |  |  |
| White | 155 (86.1%) | 78 (86.7%) | 77 (85.6%) | >0.99 |
| Non-white<sup>B</sup> | 11 (6.1%) | 6 (6.7%) | 5 (5.6%) |  |
| **Ethnicity** |  |  |  |  |
| Non-Hispanic or Latino | 168 (93.3%) | 83 (92.2%) | 85 (94.4%) | 0.33 |
| Hispanic or Latino | 8 (4.4%) | 6 (6.7%) | 2 (2.2%) |  |
| **Marital status** |  |  |  |  |
| Married | 118 (65.6%) | 69 (76.7%) | 49 (54.4%) | <0.01 |
| Unmarried | 60 (33.3%) | 21 (23.3%) | 39 (43.3%) |  |
| **Smoking status** |  |  |  |  |
| Never smoker | 76 (42.2%) | 48 (53.3%) | 28 (31.1%) | <0.01 |
| Quit | 79 (43.9%) | 36 (40.0%) | 43 (47.8%) |  |
| Current smoker | 25 (13.9%) | 6 (6.7%) | 19 (21.1%) |  |
| **BMI at first MST screening** | 25.9 (6.7) | 26.6 (7.0) | 25.5 (6.4) | 0.08 |
| **Cancer type** |  |  |  |  |
| Gastrointestinal | 42 (23.3%) | 21 (23.3%) | 21 (23.3%) | >0.99 |
| Head and neck | 88 (48.9%) | 44 (48.9%) | 44 (48.9%) |  |
| Lung | 50 (27.8%) | 25 (27.8%) | 25 (27.8%) |  |
| **Cancer stage** |  |  |  |  |
| 0/I | 42 (23.3%) | 28 (31.1%) | 14 (15.6%) | 0.09 |
| II | 31 (17.2%) | 13 (14.4%) | 18 (20.0%) |  |
| III | 30 (16.7%) | 15 (16.7%) | 15 (16.7%) |  |
| IV | 58 (32.2%) | 25 (27.8%) | 33 (36.7%) |  |
| **Surgery** |  |  |  |  |
| No | 58 (32.2%) | 26 (28.9%) | 32 (35.6%) | 0.43 |
| Yes | 122 (67.8%) | 64 (71.1%) | 58 (64.4%) |  |
| **Neoadjuvant or adjuvant chemotherapy** |  |  |  |  |
| No | 89 (49.4%) | 48 (53.3%) | 41 (45.6%) | 0.37 |
| Yes | 91 (50.6%) | 42 (46.7%) | 49 (54.4%) |  |
| **Neoadjuvant or adjuvant Radiation** |  |  |  |  |
| No | 110 (61.1%) | 59 (65.6%) | 51 (56.7%) | 0.29 |
| Yes | 70 (38.9%) | 31 (34.4%) | 39 (43.3%) |  |
| **Immunotherapy** |  |  |  |  |
| No | 152 (84.4%) | 80 (88.9%) | 72 (80.0%) | 0.15 |
| Yes | 28 (15.6%) | 10 (11.1%) | 18 (20.0%) |  |
| **Number of freeze/thaws** | 0 (0) | 0 (0) | 0 (0) | 0.75 |
| **Anticoagulation treatment** |  |  |  |  |
| ACD-Solution A | 4 (2.2%) | 2 (2.2%) | 2 (2.2%) | >0.99 |
| CPT | 14 (7.8%) | 7 (7.8%) | 7 (7.8%) |  |
| CPT, EDTA-K2 | 6 (3.3%) | 3 (3.3%) | 3 (3.3%) |  |
| EDTA-K2 | 134 (74.4%) | 67 (74.4%) | 67 (74.4%) |  |
| Sodium Citrate | 22 (12.2%) | 11 (12.2%) | 11 (12.2%) |  |
| **Days from MST score to sample collection** | 0 (28.3) | 0 (29.5) | 0 (26.8) | 0.35 |
| **GDF15 level (pg/mL)** | 1221.1<br>(773.8,<br>2043.1) | 1008.4<br>(618.6,<br>1441.8) | 1379.7<br>(931.2,<br>2513.7) | <0.001 |

Percentages may not add up to 100% due to missing data
<sup>A</sup> P-values calculated using the Mann-Whitney U Test for continuous variables and Chi Square Test of Independence for categorical variables. Fisher's Exact Test was applied to ethnicity and anticoagulation treatment due to cell counts below five.
<sup>B</sup> The non-White group includes Black or African American, Asian, American Indian or Alaska Native, Native Hawaiian or other Pacific Islander, and other race groups.
These groups were combined due to the small number of non-White participants to ensure model convergence.

## Lipid metabolites associated with malnutrition risk

Thirteen lipids were selected by elastic net regression in more than 90% of the 50 iterations (**Figure 1**), including species from various lipid classes: a cholesterol ester (**CE**), a ceramide with an 18:2;O2 sphingoid base (81), a dihexosylceramide, a sulfated hexosylceramide (sulfatide) (**SHexCer**) (82, 83), two phosphatidylcholines (**PC**), an LPC, a lysophosphatidylinositol (**LPI**), three phosphatidylethanolamines (**PE**), a sphingomyelin (**SM**), and a triacylglycerol (**TG**). In conditional logistic regression, twelve of these lipids had an inverse association with malnutrition risk, whereas dihexosylceramide 18:1;O2/20:0, was positively associated with malnutrition risk.

**Figure 1.**
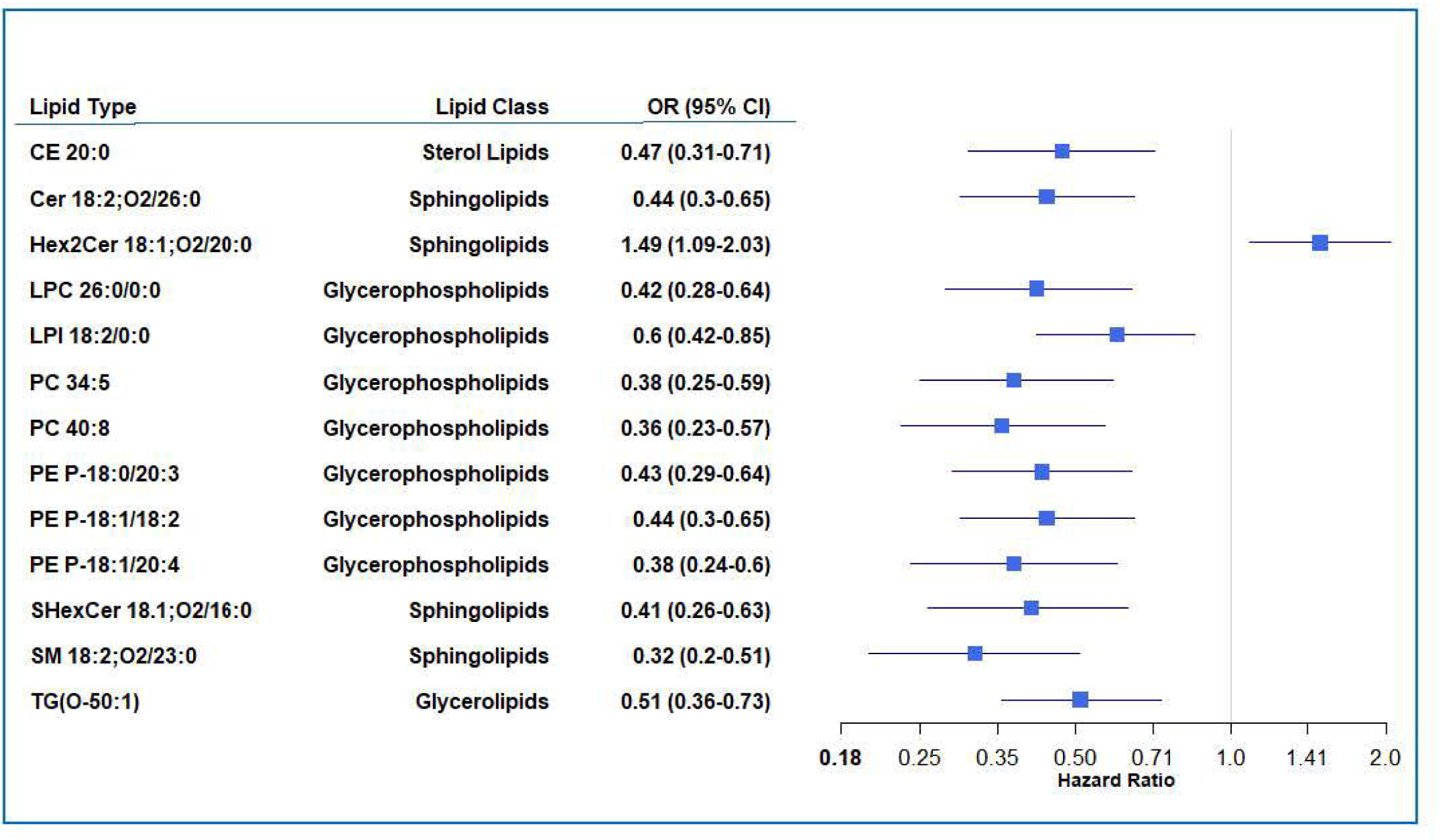
Forest plot depicting unadjusted associations with malnutrition risk (Malnutrition Screening Tool score = 0 versus ž2) for the 13 lipids consistently selected (>9O% of iterations) by elastic net regression offset for potential confounders ^A^ among 180 participants with head and neck, gastrointestinal, or lung cancers treated at the Huntsman Cancer Institute (HCI).

Absolute median (IQR) levels for all plasma lipid metabolites for cases and controls are available in **Supplementary Table 1**. Approximately 90% of the 701 lipids species analyzed through lipidomics were lower in cases compared to controls, and there was a negative mean percent difference for majority of lipid species. Total lipid levels were significantly reduced in cases compared to controls (**Figure 2**).

**Figure 2.**
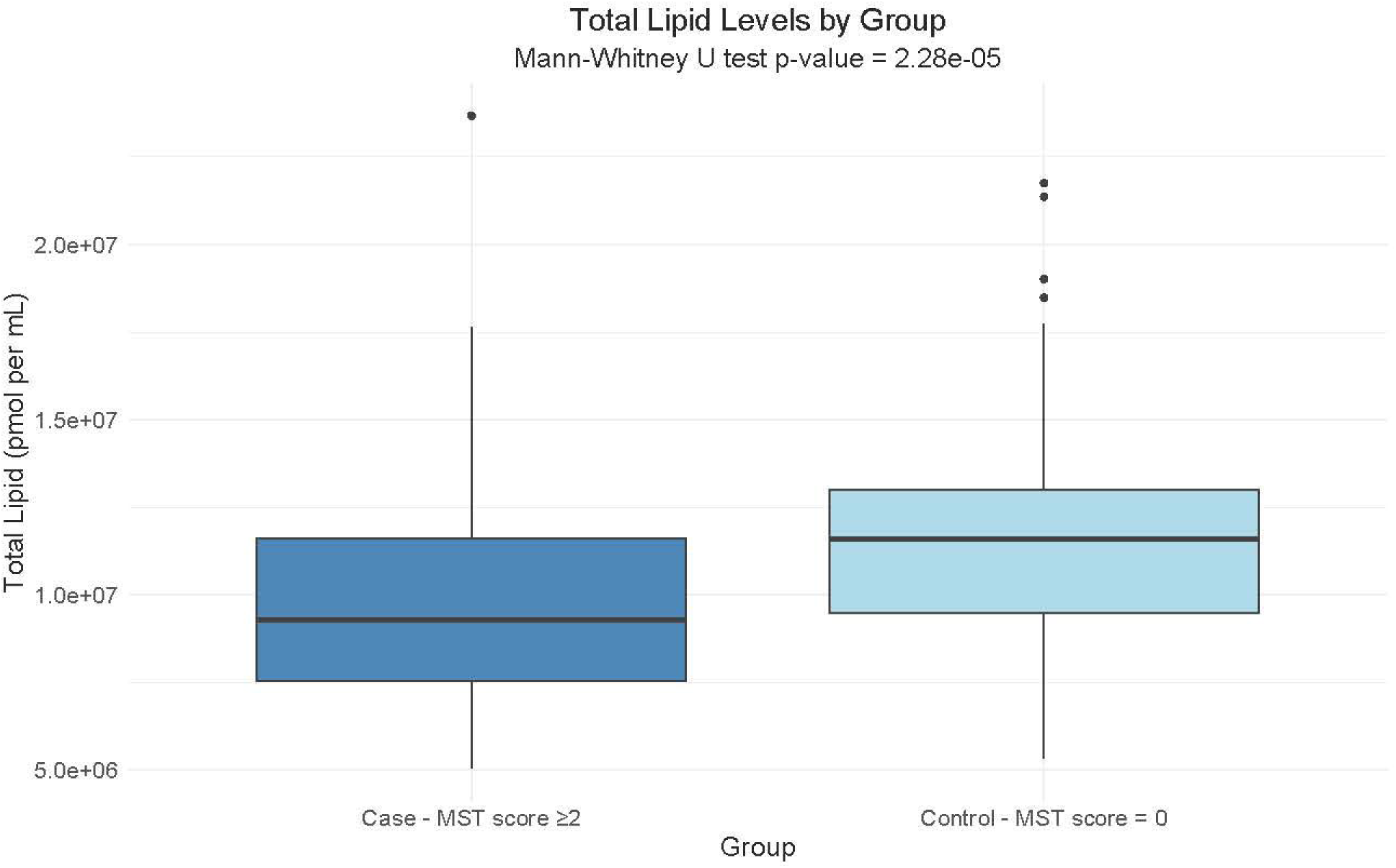
Box plot depicting total lipid levels (pmol per ml_) of the 701 analyzed lipids among 180 participants with head and neck, gastrointestinal, or lung cancers treated at the Huntsman Cancer Institute (HCI). Participants were categorized by malnutrition risk status using the Malnutrition Screening Tool (MST), with cases defined as an MST score >2 and controls as MST score = 0.

## Association of a Lipid Malnutrition Risk Score with malnutrition risk

A one-standard deviation increase in the weighted Lipid Malnutrition Risk Score, comprising the thirteen lipids selected by elastic net regression, was significantly associated with increased likelihood of malnutrition risk in both minimally (OR = 3.41, 95% CI 2.05-5.65, p < 0.001) and fully adjusted models (OR = 3.57, 95% CI 1.97-6.47, p < 0.001) (**Table 2**). In exploratory analysis, we evaluated differences by cancer type and stage (**Table 2)**. The lipid score was significantly associated with increased likelihood of malnutrition risk among patients with lung and head and neck cancers, but not gastrointestinal cancer. Fully adjusted models could not be conducted for any cancer types due to inadequate sample sizes. Additionally, the lipids score was associated with significantly increased risk for malnutrition risk among patients with stage III or IV but not stage 0/I or II cancer.

**Table 2.**
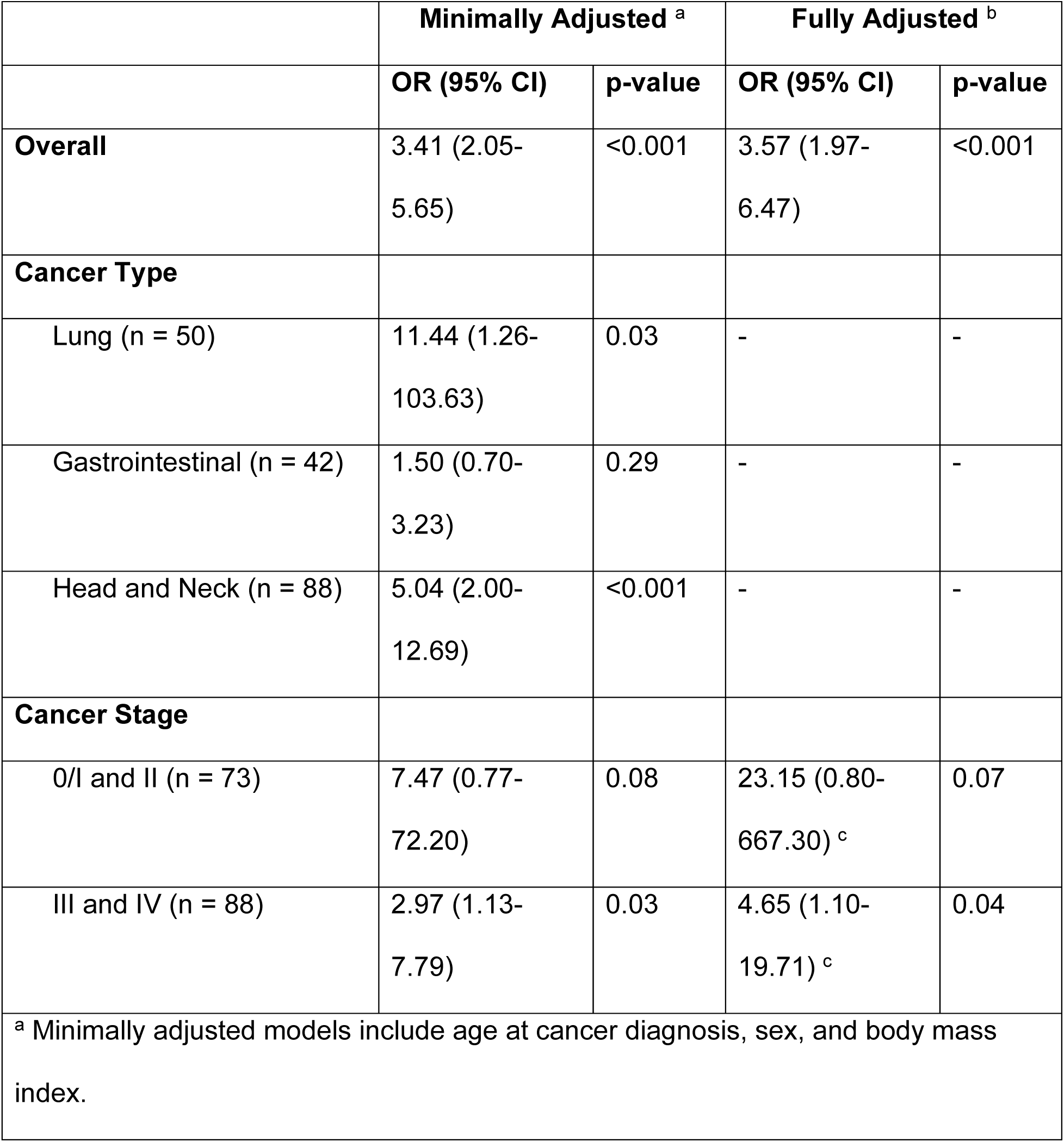
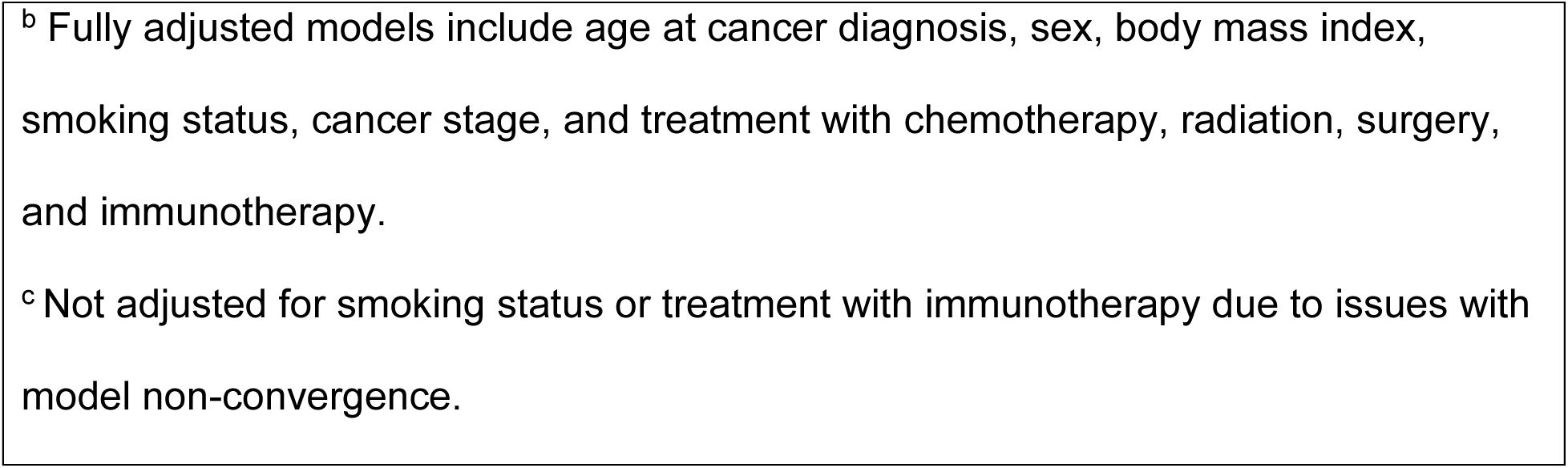
Minimally and fully adjusted associations between a one standard deviation increase in Lipid Malnutrition Risk Score with malnutrition risk (Malnutrition Screening Tool score = 0 versus ≥2) and all-cause mortality among 180 participants with head and neck, gastrointestinal, or lung cancers treated at the Huntsman Cancer Institute (HCI).

|  | Minimally Adjusted <sup>a</sup> |  | Fully Adjusted <sup>b</sup> |  |
| --- | --- | --- | --- | --- |
|  | OR (95% CI) | p-value | OR (95% CI) | p-value |
| **Overall** | 3.41 (2.05-5.65) | <0.001 | 3.57 (1.97-6.47) | <0.001 |
| **Cancer Type** |  |  |  |  |
| Lung (n = 50) | 11.44 (1.26-103.63) | 0.03 | - | - |
| Gastrointestinal (n = 42) | 1.50 (0.70-3.23) | 0.29 | - | - |
| Head and Neck (n = 88) | 5.04 (2.00-12.69) | <0.001 | - | - |
| **Cancer Stage** |  |  |  |  |
| 0/I and II (n = 73) | 7.47 (0.77-72.20) | 0.08 | 23.15 (0.80-667.30) <sup>c</sup> | 0.07 |
| III and IV (n = 88) | 2.97 (1.13-7.79) | 0.03 | 4.65 (1.10-19.71) <sup>c</sup> | 0.04 |
| <sup>a</sup> Minimally adjusted models include age at cancer diagnosis, sex, and body mass index. |  |  |  |  |

<sup>b</sup> Fully adjusted models include age at cancer diagnosis, sex, body mass index, smoking status, cancer stage, and treatment with chemotherapy, radiation, surgery, and immunotherapy.
<sup>c</sup> Not adjusted for smoking status or treatment with immunotherapy due to issues with model non-convergence.

Analysis of the lipid score’s predictive performance using ROC-AUC C-statistic found that the addition of the Lipid Malnutrition Risk Score to a model containing established malnutrition risk factors increased the C-statistic from 0.78 (95% CI 0.71- 0.84) to 0.90 (95% CI 0.86-0.94) (**Figure 3**). A model with the lipid score alone performed slightly better than risk factors alone (C statistic = 0.85, 95% CI 0.79-0.91). Incorporation of the lipid score also resulted in significant improvements in continuous NRI (NRI = 1.07, 95% CI 0.82-1.31), and IDI (IDI = 0.20, 95% CI 0.16-0.25) when compared to risk factors alone (**Supplementary Table 2**).

**Figure 3.**
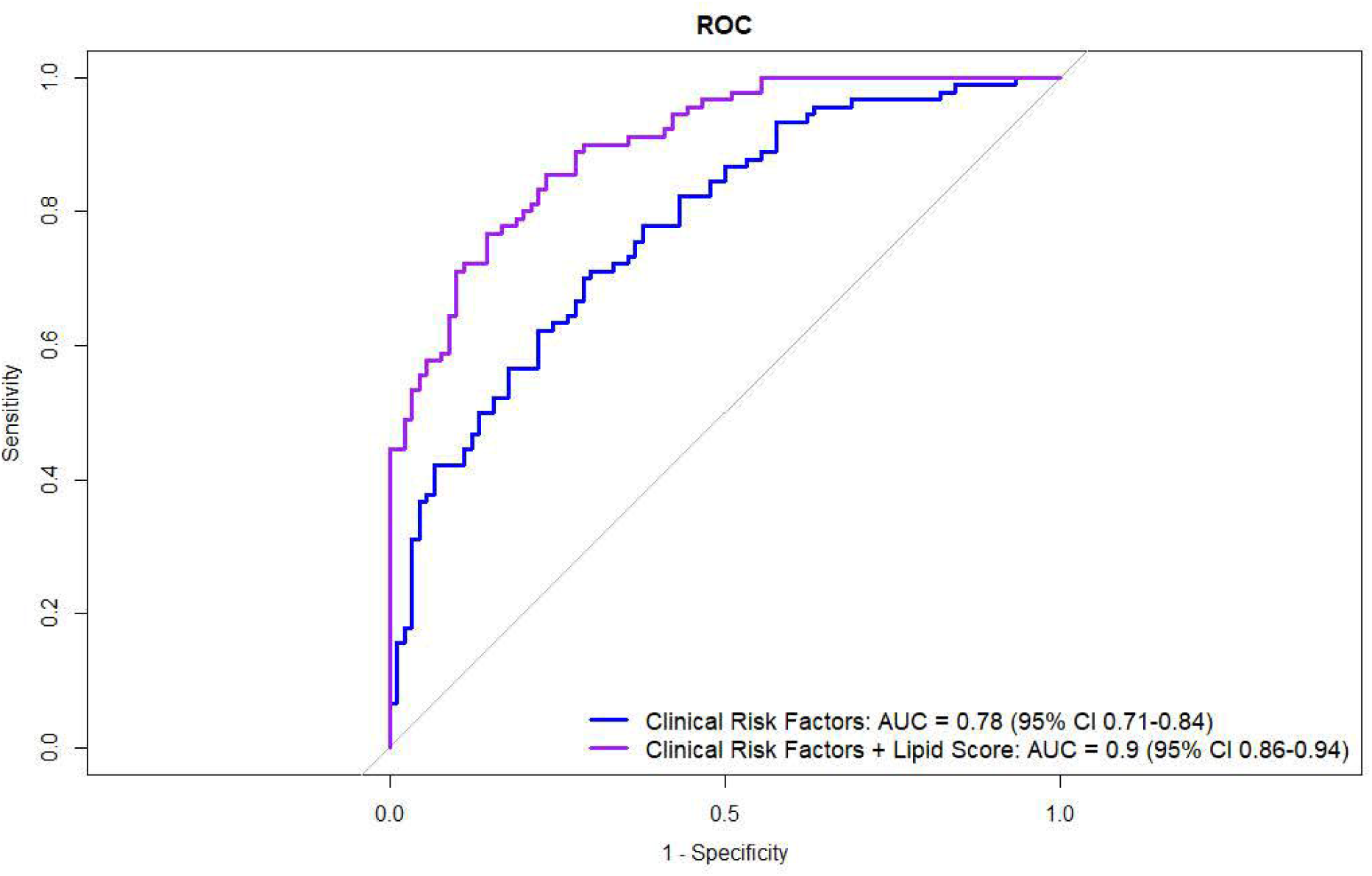
Receiver Operating Characteristic Curves **(ROC-AUC)** for conditional logistic regression models comparing clinical risk factors for malnutrition alone (age at cancer diagnosis, sex, body mass index, smoking status, treatment with chemotherapy, radiation therapy, surgery, and immunotherapy) and combined with the Lipid Malnutrition Risk Score among 180 participants with head and neck, gastrointestinal, or lung cancers treated at the Huntsman Cancer Institute **(HCI).**

## Enrichment of lipid classes and properties by malnutrition risk status

A bubble plot displaying lipid terms significantly enriched in the lipid enrichment analysis of plasma lipidomic data is presented in **Figure 4**. The top five enriched lipid classes and biophysical properties were: headgroup with positive charge/zwitter ion, glycerophosphocholines, endoplasmic reticulum, glycerolipids, and diacylglycerophosphoethanolamines. Lipid terms associated with membrane structure and signaling (e.g., glycerophospholipids) were significantly downregulated in cases compared to controls, whereas lipid terms related to storage (e.g., glycerolipids) were upregulated in patients with MST score ≥2 (**Figure 5).** A full summary of up- and downregulated lipid terms is available in **Supplementary Table 3.**

**Figure 4.**
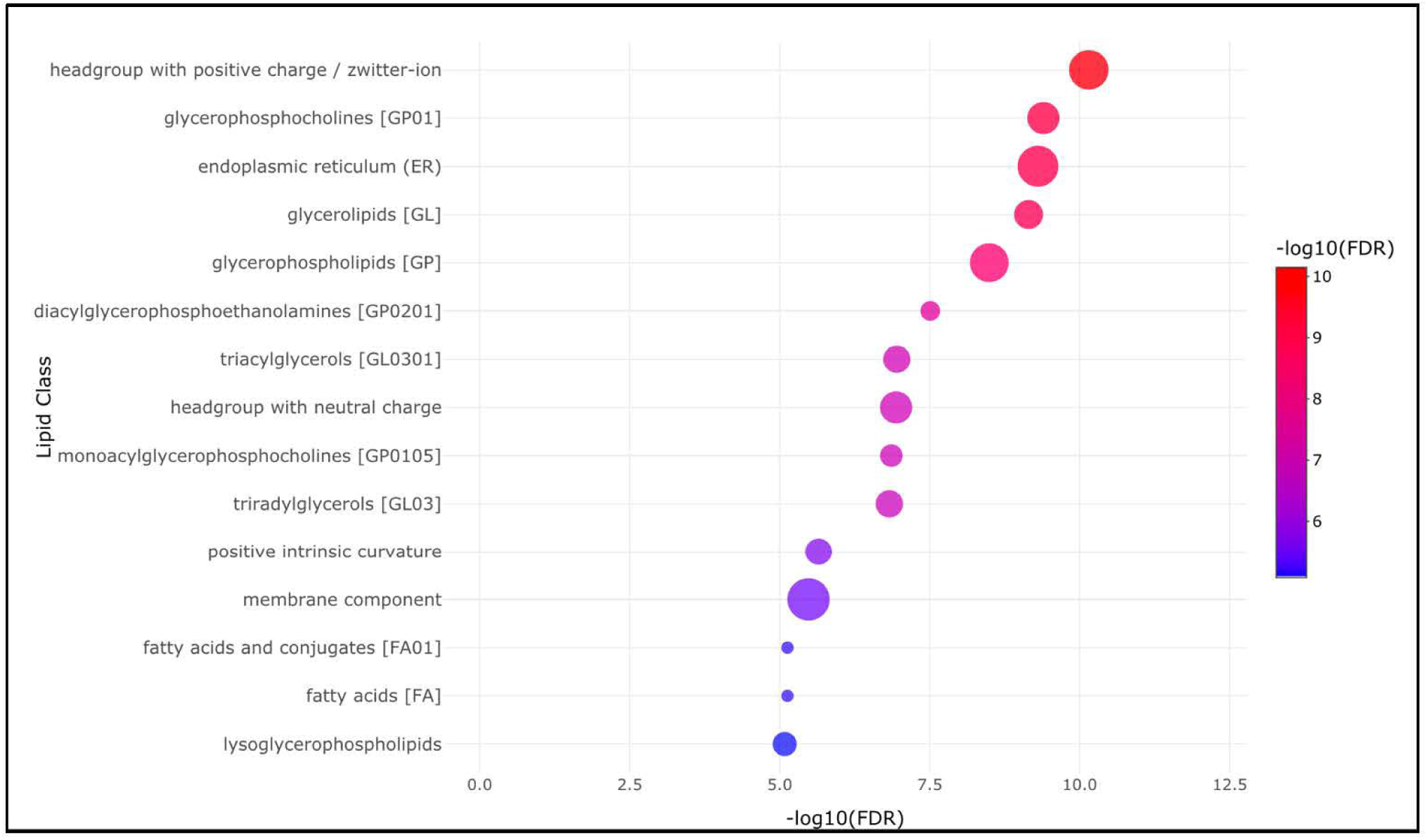
Bubble plot illustrating the top 15 significantly enriched lipid ontology terms (FDR < 0.05) identified through Lipid Ontology (LION) enrichment analysis using plasma lipidomics data from 180 participants with head and neck, gastrointestinal, or lung cancers treated at the Huntsman Cancer Institute **(HCI).** Participants were categorized by malnutrition risk status using the Malnutrition Screening Tool (MST), with cases defined as an MST score >2 and controls as MST score = 0. Bubble size denotes the number of annotated lipids in each LION term.

**Figure 5.**
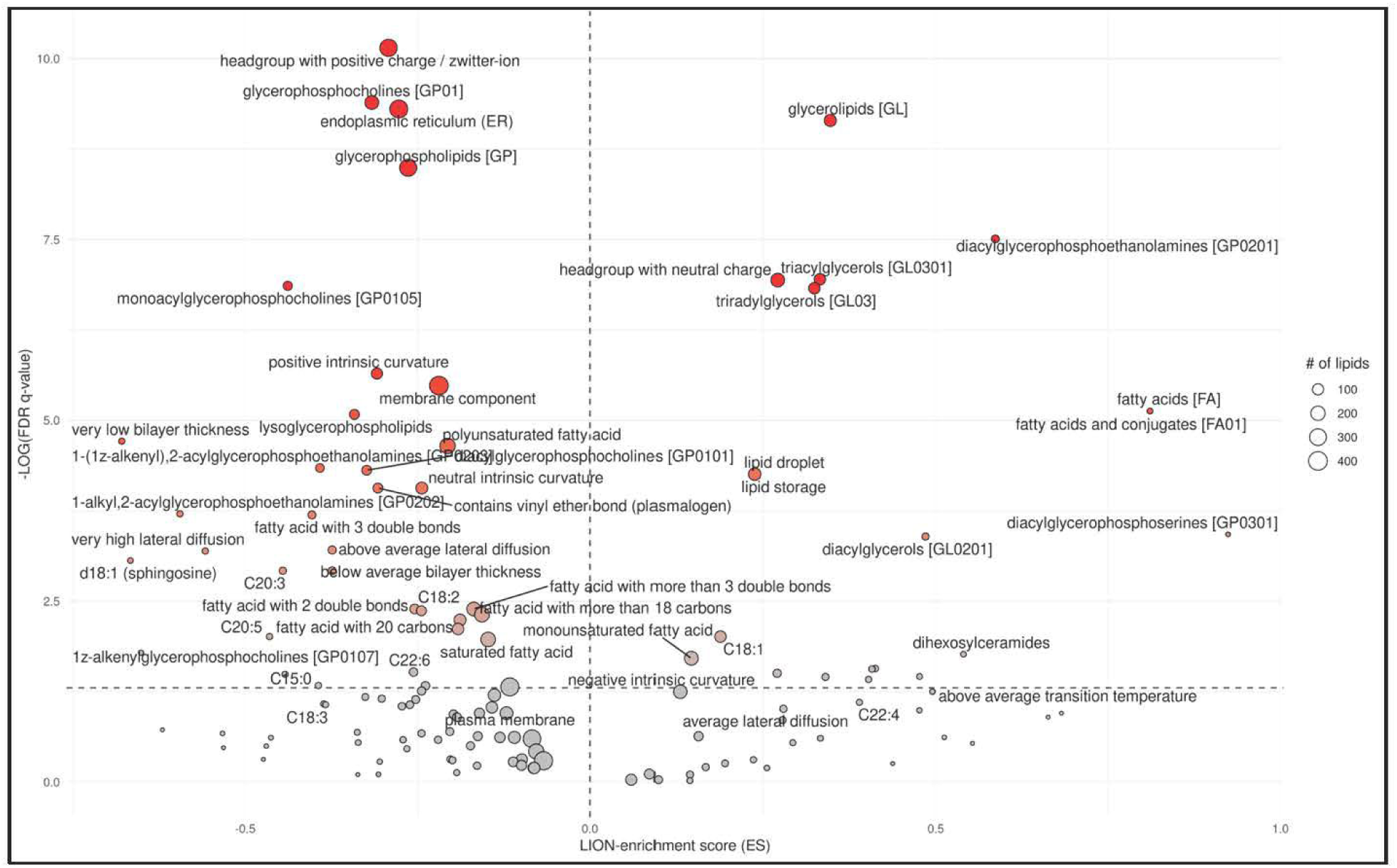
Volcano plot depicting differentially enriched lipid terms identified through Lipid Ontology (LION) enrichment analysis using plasma lipidomics data from 180 participants with head and neck, gastrointestinal, or lung cancers treated at the Huntsman Cancer Institute (HCI). Participants were categorized by malnutrition risk status using the Malnutrition Screening Tool (MST), with cases defined as an MST score >2 and controls as MST score = 0. Gray nodes represent LION-terms with no or weak enrichment, while red nodes indicate strongly enriched terms. The horizontal gray line marks the statistical significance threshold (q < 0.05, corresponding to -logw(q) >1.3)

## Sensitivity Analyses

After adjustment for participants’ GDF15 levels, thirteen lipids were selected by elastic net regression in >90% of the 50 iterations (**Supplementary Table 4**). PE P- 18:1/20:4, PE P-18:0/20:3, and ceramide 18:2;O2/26:0 were no longer retained. Three new lipids were selected: PC P-18:0/18:2, a TG representing an alternative annotation of the TG identified in primary analysis (TG(O-50:1)[SIM]), and one additional triacylglycerol. The TG(O-50:1)[SIM] annotation captures all TG(O-50:1) species regardless of acyl chain composition, whereas the NL-18:1 version specifically reflects those containing an acyl 18:1 chain. All three newly selected lipid species were inversely associated with malnutrition risk. In the second sensitivity analysis, additional adjustment for imperfectly matched matching factors had minimal effects on selected lipids (**Supplementary Table 5**). In the final sensitivity analysis excluding patients with a cancer diagnosis before 2021, the year of MST implementation at HCI outpatient clinics, 11 lipids were selected by elastic net (**Supplementary Table 6**). The lipid species selected were broadly consistent with those identified in the primary analysis.

## Discussion

This study identified 13 lipids associated with malnutrition risk using elastic net regression. A weighted Lipid Malnutrition Risk Score derived from these lipids was associated with approximately a threefold increase in the odds of malnutrition risk. The association of the risk score remained positively associated with malnutrition risk when stratified by cancer type, though results for gastrointestinal cancers did not reach statistical significance, likely due to small sample size. Moreover, the lipid score improved the prediction of malnutrition risk status beyond known risk factors, increasing the ROC-AUC C-statistic from 0.78 to 0.90. Lipid enrichment analysis further revealed down-regulation of lipid classes and properties associated with membrane structure and signaling in individuals with malnutrition risk and upregulation of those related to lipid storage.

Early identification of oncology patients at risk for malnutrition using validated screening tools is recommended to facilitate timely referral to RDs for comprehensive nutritional assessment and subsequent nutrition intervention to improve patient prognosis (21). However, no robust malnutrition biomarkers have been identified with potential to enhance existing screening tools, which rely on patient-reported subjective information, are prone to misclassification, and place additional demands on healthcare professionals operating under significant time constraints (32). Lipidomics and metabolomics hold potential for identification of novel biomarkers of malnutrition status (33–36).

Few prior studies have investigated circulating metabolites as potential markers of malnutrition (41). One study conducted among 418 patients with gastric cancer identified 25 metabolites and lipids that were differentially expressed between individuals with and without malnutrition (41). Consistent with the findings of this analysis, malnourished patients had reduced levels of LPCs, and glycerophospholipid metabolism was among the top eight enriched pathways (41). LPCs are membrane lytic lipids produced via the enzymatic cleavage of PCs, which are key components of the plasma membrane (84, 85). In this study, we observed reductions in both LPCs and PCs species. As LPCs are derived from PCs, reduction in LPCs often reflects underlying decreases in PCs (86).

Low plasma LPC levels have been associated with impaired muscle mitochondrial function and decreased muscle quality (86, 87). Mechanistic studies in mice have also documented a central role of PCs in dietary lipid absorption (88), and LPC supplementation has been reported to facilitate dietary fat absorption in the gut to mitigate fat malabsorption (89, 90). In this analysis, total serum lipid concentrations were lower in patients at risk for malnutrition compared to those without risk, which could further suggest impaired lipid absorption. However, insufficient dietary intake, reduced body weight, or altered lipid metabolism in patients with malnutrition risk may also play a role. Additionally, low plasma LPC levels inversely correlate with inflammatory markers such as C-reactive protein (**CRP**) (85) and are reduced in patients with inflammatory illnesses, such as pneumonia and sepsis (91–93), suggesting inflammation could underlie the association between LPCs and malnutrition.

Decreased plasma LPCs have also been reported in patients with cancer-related cachexia (45, 46, 94–97), a condition defined by involuntary loss of body weight and muscle mass, with or without fat loss, which is thought to be driven by inflammation and metabolic dysregulation (98). Cachexia and malnutrition share overlapping clinical features and treatment approaches (28, 29, 99, 100). However, cachexia, particularly in its later stages, cannot be fully reversed by nutrition support (99). The similarities between these conditions, including the presence of unintentional weight loss, reduced food intake, and loss of muscle and adipose tissue, may contribute to comparable shifts in lipids species. Indeed, adjustment for GDF15 levels in our sensitivity analysis had minimal impact on findings, suggesting that these biomarkers may extend to informing cachexia risk.

LION analysis indicated that neutral lipids involved in energy storage, such as TGs and diacylglycerols (**DGs**), were elevated in the plasma of patients at risk for malnutrition compared to those with no risk. The overall increase in storage-related lipids may be related to the presence of underlying cancer-associated cachexia and inflammation (98). As previously noted, malnutrition and cachexia share many defining features and can co-exist; therefore, patients identified as at risk by MST could be experiencing both conditions concurrently. In cachexia, inflammation and resulting insulin resistance along with heightened sensitivity to lipolytic factors drive elevated levels of lipolysis, leading to increased flux of free fatty acids into circulation (101, 102). These fatty acids are taken up by the liver, re-esterified into TGs, and secreted as very low-density lipoproteins (**VLDL**). These metabolic disturbances are hypothesized to contribute not only to increased production of VLDL but also to impaired VLDL clearance, collectively resulting in elevated TG levels (101, 103). Yet, findings on whether TG levels are increased or decreased in the context of malnutrition or cachexia have been inconsistent (104–106). These discrepancies may be attributable to differences in disease duration, severity, or cancer type. While enrichment analysis highlights overall class-level lipid changes, individual lipids within a class may deviate from these broader trends. For example, although LION analysis showed an overall increase in storage lipids, specific ether-linked TG species, like TG (O-51:1), were downregulated. This may reflect reduced susceptibility to lipolysis, as key enzymes such as Hormone Sensitive Lipase and Adipose Triacylglycerol Lipase primarily target ester bonds (107).

Our analysis also identified several sphingolipids associated with malnutrition risk. As essential constituents of plasma membranes, sphingolipids contribute to signaling and structural organization of membrane-bound receptors (108). Dysregulated sphingolipid metabolism has been observed in states of both under- and overnutrition (39, 45, 96, 109), and elevated levels of sphingolipids, particularly ceramides, have been reported in patients with cancer-associated cachexia (45, 96). Increased ceramide levels may result from heightened activity of secretory sphingomyelinase in response to inflammation, which promotes the conversion of sphingomyelins to ceramides (110, 111). Consistent with these findings, our lipid enrichment analysis showed that ceramides and dihexosylceramides were collectively upregulated in patients at risk for malnutrition, whereas sphingomyelins were downregulated. Conversely, elastic net identified an individual ceramide, 18:2;O2/26:0, that was inversely associated with malnutrition risk. This ceramide contains a sphingadiene base, distinguishing it from canonical ceramides, which are built on a sphingosine backbone (112). The unique structure of sphingadiene-based ceramides is hypothesized to confer distinct membrane properties and biologic functions (112–114). However, despite emerging interest, the biologic roles of this sphingolipid subclass are not well defined. Similarly, we identified a sulfated hexosylceramide, SHexCer 18:2;O2/23:0, that was inversely associated with malnutrition risk. Altered levels of sulfated hexosylceramides, also known as sulfatides, have been linked to cognitive disorders such as Alzheimer’s (115–117) and to several cancers (118–121), but to our knowledge, have not previously been associated with malnutrition or cancer-associated cachexia. Further research is needed to confirm the role of this sphingolipid species in malnutrition and to identify potential underlying mechanisms.

Strengths of this analysis include use of a validated malnutrition screening tool to define malnutrition risk, plasma collected within ±30 days of an MST score, and inclusion of three different cancer types. This analysis utilized the University of Utah’s Metabolomic and Proteomics Core to ensure valid and reliable quantification of GDF15 and lipid species. The study has limitations. Exposure classification relied on a malnutrition screening tool rather than comprehensive malnutrition assessment due to current clinic practices. Additionally, residual confounding is possible as we were limited to adjusting for covariates that were available in the EMR. While internal validation was used for elastic net regression analyses, we could not conduct external validation. LC-MS methodology does not allow for precise determination of lipid structural features such as double bond position. For example, in ceramide 18:2;O2/26:0, the location of the second double bond is ambiguous. We assumed the (4E,14Z) configuration, as it is the most frequently observed form in mammalian tissue (114, 122). We leveraged plasma GDF15 levels as a proxy for cachexia. However, it’s plausible that GDF15 levels may not have fully captured the presence or severity of cachexia, potentially resulting in residual confounding. Given the metabolic heterogeneity among tumors from different tissues, the utility of a single lipidomic panel is uncertain and warrants further investigation (123). We were unable to match for fasting status as this information was not collected for plasma samples, and the population included in this analysis was primarily white, limiting generalizability. Finally, while lipidomics and metabolomics show promise for identifying biomarkers of malnutrition risk, the high cost and limited accessibility of these analyses in standard care settings may hinder their integration into routine clinical practice.

## Conclusion

There are no established biomarkers for malnutrition screening. This study highlights the potential for lipidomics to identify circulating biomarkers of malnutrition risk and suggests lipid classes and biophysical properties that may be differentially regulated in individuals with and without malnutrition risk. Large, prospective studies in diverse populations are needed to validate and expand upon these findings.

## Data Availability

The data described in this manuscript and analytic code will be made available from the corresponding author upon reasonable request.

## Acknowledgements

We acknowledge the Total Cancer Care (TCC) study participants at Huntsman Cancer Institute for their participation and contributions to this work. We also acknowledge the valuable contributions of the Biorepository and Molecular Pathology (BMP) Shared Resource, the University of Utah Drug Discovery Core, the TCC disease center leaders, and the University of Utah Metabolomics Core. Research reported in this publication utilized the Research Informatics Shared Resource at Huntsman Cancer Institute at the University of Utah and was supported by the National Cancer Institute of the National Institutes of Health under Award Number P30CA042014. The content is solely the responsibility of the authors and does not necessarily represent the official views of the NIH.

## Authorship Contributions

Conceptualization, M.C.P., R.H.; Investigation, J.E.C., J.A.M., B.L., P.S.; Data Curation, J.A.M., B.L.; Formal analysis, M.C.P., R.H.; Validation, M.C.P.; Writing – original draft preparation, R.H., M.C.P; Writing – review and editing, all authors; Visualization, R.H., P.S.; Supervision, M.C.P., K.W., A.M.C., A.C., A.S.; Funding acquisition, M.C.P. All authors have read and agreed to the final version of the manuscript.

## Declaration of Generative AI and AI-Assisted Technology in the Writing Process

During the preparation of this work the author used ChatGPT and Microsoft Copilot for editing assistance. After using these tools, the author reviewed and edited the content as needed and takes full responsibility for the content of this publication

## Abbreviations

**ACD**, Acid Citrate Dextrose; **BMI**, Body Mass Index; **CE**, Cholesteryl Ester; **CPT**, Cell Preparation Tube; **DG**, Diacylglycerol; **EDTA**, Ethylenediaminetetraacetic Acid; **EMR**, Electronic Medical Record; **ELISA**, Enzyme-Linked Immunosorbent Assay; **FDR**, False Discovery Rate; **GDF15**, Growth/Differentiation Factor 15; **HCI**, Huntsman Cancer Institute; **ICD-O**, International Classification of Diseases for Oncology; **IDI**, Integrated Discrimination Improvement; **IRB**, Institutional Review Board; **LC-MS**, Liquid Chromatography-Mass Spectrometry; **LC-MS/MS**, Liquid Chromatography-Tandem Mass Spectrometry; **LION**, Lipid Ontology; **LPC**, Lysophosphatidylcholine; **LPI**, Lysophosphatidylinositol; **MST**, Malnutrition Screening Tool; **NRI**, Net Reclassification Improvement; **OR**, Odds Ratio; **PC**, Phosphatidylcholine; **PE**, Phosphatidylethanolamine; **PMM**, Predictive Mean Matching; **QC**, Quality Control; **RD**, Registered Dietitian; **ROC-AUC**, Receiver Operating Characteristic - Area Under the Curve; **SERRF**, Systematic Error Removal using Random Forest; **SHexCer**, Sulfated Hexosylceramide; **SM**, Sphingomyelin; **TCC**, Total Cancer Care; **TG**, Triacylglycerol; **VLDL**, Very Low-Density Lipoprotein.

