## Supplementary Material for "Identification of circulating lipidomic biomarkers of malnutrition risk among oncology patients in the Total Cancer Care (TCC) Study: a cross-sectional analysis"

**Supplementary Table 1.** Absolute plasma lipid metabolite levels for the 701 lipids included in lipidomics analysis sorted by down or upregulation in cases compared to controls and false detection rate (FDR) adjusted p-value. Cases are defined as a malnutrition screening tool (MST) score ≥2 and controls are defined as MST score = 0. Patients had lung, gastrointestinal, or head and neck cancer and were receiving treatment at the Huntsman Cancer Institute (HCI).

| Lipid (pmol per mL) | Direction | Mean % difference ^a^ | Case median | Control median | FDR adjusted p-value ^b^ |
| --- | --- | --- | --- | --- | --- |
| SM 18:2;O2/23:0 | DOWN | -30.11 | 2994.06 | 4278.84 | <0.0001 |
| Cer 16:1;O2/24:0 | DOWN | -43.54 | 270.13 | 476.86 | <0.0001 |
| Cer 18:2;O2/23:0 | DOWN | -36.10 | 209.52 | 349.87 | <0.0001 |
| PC 14:0_20:4 | DOWN | -40.91 | 747.96 | 1540.56 | <0.0001 |
| PC 34:5 | DOWN | -53.15 | 34.81 | 87.56 | <0.0001 |
| Cer 18:2;O2/24:0 | DOWN | -36.72 | 522.61 | 893.41 | <0.0001 |
| PC 14:0_22:6 | DOWN | -39.91 | 180.15 | 312.57 | <0.0001 |
| SM 16:1;O2/23:0 \| SM 17:1;O2/22:0 | DOWN | -29.67 | 3263.70 | 4583.64 | <0.0001 |
| PC 38:6 a | DOWN | -30.70 | 4463.38 | 7149.90 | <0.0001 |
| Cer 16:1;O2/23:0 | DOWN | -40.66 | 86.90 | 147.14 | <0.0001 |
| Cer 17:1;O2/24:0 | DOWN | -35.35 | 174.36 | 283.92 | <0.0001 |
| PC 40:8 | DOWN | -24.36 | 482.56 | 669.49 | <0.0001 |
| Cer 17:1;O2/23:0 | DOWN | -32.01 | 53.47 | 81.53 | <0.0001 |
| PC 32:2 | DOWN | -39.36 | 2760.97 | 4634.48 | <0.0001 |
| SM 18:2;O2/22:0 | DOWN | -23.09 | 9289.63 | 12334.45 | <0.0001 |
| SM 18:2;O2/14:0 | DOWN | -26.34 | 689.77 | 1054.67 | <0.0001 |
| SM 41:0;O2 | DOWN | -25.05 | 1353.77 | 1851.33 | <0.0001 |
| SM 18:1;O2/23:0 \| SM 17:1;O2/24:0 | DOWN | -25.05 | 7642.33 | 10792.97 | <0.0001 |
| PC 18:2_20:5 | DOWN | -45.05 | 190.50 | 315.27 | <0.0001 |
| LPC 14:0 sn2 | DOWN | -34.40 | 218.65 | 364.24 | <0.0001 |
| CE 18:3 | DOWN | -32.62 | 231305.46 | 380834.95 | <0.0001 |
| LPC 22:0 sn1 | DOWN | -30.76 | 18.77 | 29.16 | <0.0001 |
| PE O-16:0/22:6 | DOWN | -28.83 | 356.03 | 534.13 | <0.0001 |
| PE O-38:5 a | DOWN | -26.78 | 328.74 | 510.27 | <0.0001 |
| PE P-16:0/22:5 n3 | DOWN | -20.56 | 1373.52 | 2094.97 | <0.0001 |
| Cer 18:1;O2/24:0 | DOWN | -27.60 | 3025.08 | 4189.05 | <0.0001 |
| LPC 14:0 sn1 | DOWN | -32.89 | 844.55 | 1332.68 | <0.0001 |
| PE P-18:1/20:4 a | DOWN | -23.75 | 1361.20 | 2280.49 | <0.0001 |
| PC 35:5 | DOWN | -36.41 | 46.16 | 93.44 | <0.0001 |
| PE P-18:0/20:3 b | DOWN | -30.53 | 36.44 | 56.19 | <0.0001 |
| PC 36:6 a | DOWN | -39.91 | 102.15 | 195.06 | <0.0001 |
| PC 40:7 a | DOWN | -20.69 | 470.86 | 667.33 | <0.0001 |
| PC 18:2/18:2 | DOWN | -36.64 | 12756.85 | 20632.39 | <0.0001 |
| PE O-18:0/22:5 | DOWN | -23.95 | 84.02 | 128.30 | <0.0001 |
| PE P-16:0/18:2 | DOWN | -28.48 | 829.64 | 1584.50 | <0.0001 |
| LPC 24:0 sn1 | DOWN | -24.84 | 44.64 | 64.54 | <0.0001 |
| PE P-18:1/18:2 b | DOWN | -35.48 | 81.47 | 152.94 | <0.0001 |
| PE P-18:0/18:2 | DOWN | -26.15 | 705.63 | 1234.32 | <0.0001 |
| PE P-16:0/20:5 | DOWN | -32.81 | 183.66 | 366.90 | <0.0001 |
| dimethyl-CE 18:2 | DOWN | -36.94 | 4169.89 | 7987.07 | <0.0001 |
| PE P-18:1/18:2 a | DOWN | -29.49 | 344.56 | 612.79 | <0.0001 |
| PE P-18:1/20:4 b | DOWN | -26.61 | 258.58 | 435.39 | <0.0001 |
| LPC 26:0 sn1 | DOWN | -20.20 | 13.30 | 16.63 | <0.0001 |
| PC 33:2 | DOWN | -29.15 | 2253.37 | 3345.73 | <0.0001 |
| Cer 18:0;O2/24:0 | DOWN | -26.79 | 176.35 | 253.32 | <0.0001 |
| Cer 18:1;O2/23:0 | DOWN | -25.68 | 1141.85 | 1518.03 | <0.0001 |
| PE O-16:0/20:4 | DOWN | -24.68 | 289.30 | 465.30 | <0.0001 |
| PE P-18:0/20:4 | DOWN | -15.46 | 2871.88 | 4354.28 | <0.0001 |
| SM 17:1;O2/14:0 | DOWN | -27.56 | 177.17 | 283.06 | <0.0001 |
| PE P-18:1/20:5 a | DOWN | -37.01 | 80.79 | 139.01 | 0.0001 |
| LPC 18:0 sn1 | DOWN | -20.97 | 19812.86 | 26903.03 | 0.0001 |
| PE P-18:0/20:5 | DOWN | -30.67 | 151.00 | 275.55 | 0.0001 |
| Cer 18:2;O2/26:0 | DOWN | -24.31 | 11.99 | 15.77 | 0.0001 |
| LPC 19:0 sn1 b | DOWN | -25.96 | 53.50 | 79.20 | 0.0001 |
| PC 15:0_20:4 | DOWN | -22.98 | 844.90 | 1174.40 | 0.0001 |
| Cer 16:1;O2/22:0 | DOWN | -29.65 | 166.10 | 251.46 | 0.0001 |
| PE P-18:0/18:1 | DOWN | -3.53 | 434.84 | 626.84 | 0.0001 |
| PC 16:0_20:5 | DOWN | -23.97 | 7419.84 | 12886.92 | 0.0001 |
| PE O-36:5 | DOWN | -31.73 | 36.91 | 60.25 | 0.0001 |
| PE P-18:1/20:3 | DOWN | -24.59 | 65.28 | 101.62 | 0.0001 |
| CE 20:5 | DOWN | -32.82 | 100791.75 | 161234.59 | 0.0001 |
| PC P-18:0/18:2 | DOWN | -27.37 | 783.80 | 1062.89 | 0.0001 |
| PE O-16:0/18:2 | DOWN | -23.73 | 65.79 | 107.28 | 0.0001 |
| DE 18:2 | DOWN | -26.45 | 7541.61 | 11440.59 | 0.0001 |
| PC 15:0_20:3 | DOWN | -24.70 | 706.04 | 899.03 | 0.0001 |
| LPC 22:0 sn2 | DOWN | -26.59 | 7.56 | 10.10 | 0.0001 |
| LPC 20:5 sn1 | DOWN | -30.44 | 316.58 | 543.89 | 0.0001 |
| PE O-18:0/20:4 | DOWN | -19.58 | 217.58 | 354.83 | 0.0001 |
| LPC 20:5 sn2 | DOWN | -27.74 | 126.38 | 218.12 | 0.0001 |
| LPC 24:0 sn2 | DOWN | -20.08 | 22.79 | 30.56 | 0.0001 |
| PC 18:0_18:2 | DOWN | -22.49 | 113175.39 | 156449.52 | 0.0001 |
| SM 18:1;O2/14:0 \| SM 16:1;O2/16:0 | DOWN | -18.16 | 9920.07 | 12651.22 | 0.0001 |
| LPC 18:2 sn1 | DOWN | -31.35 | 19353.10 | 25173.27 | 0.0001 |
| LPC 15-MHDA sn2 | DOWN | -23.40 | 100.19 | 137.00 | 0.0001 |
| LPC 18:0 sn2 | DOWN | -19.92 | 5842.34 | 7638.26 | 0.0001 |
| methyl-CE 18:1 | DOWN | -26.68 | 2788.63 | 4617.32 | 0.0001 |
| LPC 18:3 (104) sn1 | DOWN | -31.26 | 298.43 | 458.41 | 0.0001 |
| LPC 16:0 sn1 | DOWN | -19.12 | 67432.56 | 90238.67 | 0.0001 |
| dimethyl-CE 18:1 | DOWN | -29.46 | 1595.82 | 2506.70 | 0.0001 |
| PC P-16:0/18:2 | DOWN | -22.99 | 5840.18 | 7822.56 | 0.0002 |
| PE P-18:0/22:6 | DOWN | -24.27 | 905.25 | 1291.06 | 0.0002 |
| LPC 15-MHDA sn1 \| LPC 17:0 sn2 | DOWN | -21.83 | 505.97 | 679.55 | 0.0002 |
| LPC 17:1 sn1 b | DOWN | -28.68 | 24.62 | 34.61 | 0.0002 |
| LPC 18:3 sn1 b | DOWN | -27.66 | 196.37 | 298.00 | 0.0002 |
| PE O-18:0/22:6 | DOWN | -25.93 | 150.12 | 227.60 | 0.0002 |
| SM 34:3;O2 | DOWN | -17.74 | 68.28 | 91.66 | 0.0002 |
| PC 28:0 | DOWN | -34.43 | 246.60 | 454.71 | 0.0002 |
| total_lipid | DOWN | -15.64 | 9265664.80 | 11578971.60 | 0.0002 |
| LPC 15:0 sn2 | DOWN | -23.14 | 253.01 | 316.30 | 0.0002 |
| LPC 18:2 sn2 | DOWN | -27.67 | 9308.81 | 11354.42 | 0.0002 |
| LPC 18:3 sn1 a \| LPC 18:3 sn2 b | DOWN | -30.96 | 235.55 | 350.28 | 0.0002 |
| TG(O-50:1) [NL-18:1] | DOWN | -41.88 | 65.68 | 112.33 | 0.0002 |
| CE 18:0 | DOWN | -25.28 | 20754.58 | 31030.76 | 0.0002 |
| HexCer 16:1;O2/24:0 | DOWN | -22.97 | 43.60 | 60.57 | 0.0002 |
| PC P-16:0/20:5 | DOWN | -32.78 | 227.17 | 370.96 | 0.0002 |
| PE P-18:1/18:1 b | DOWN | -6.34 | 27.25 | 40.29 | 0.0002 |
| Cer 18:2;O2/22:0 | DOWN | -22.41 | 301.21 | 446.17 | 0.0002 |
| PE O-34:1 | DOWN | -15.25 | 69.63 | 101.75 | 0.0002 |
| PC 39:5 b | DOWN | -23.48 | 129.19 | 164.12 | 0.0002 |
| PE O-18:1/22:6 | DOWN | -19.99 | 108.48 | 136.55 | 0.0002 |
| TG(O-50:1) [NL-16:0] | DOWN | -41.14 | 133.91 | 209.24 | 0.0002 |
| DE 20:4 | DOWN | -17.72 | 1968.34 | 2782.65 | 0.0002 |
| LPC 19:0 sn1 a \| LPC 19:0 sn2 b | DOWN | -21.20 | 30.84 | 41.77 | 0.0002 |
| LPC 17:0 sn1 | DOWN | -21.02 | 815.05 | 1043.67 | 0.0002 |
| TG(O-50:2) [NL-18:2] | DOWN | -39.30 | 30.84 | 50.80 | 0.0002 |
| LPC 16:0 sn2 | DOWN | -17.74 | 17251.24 | 21706.55 | 0.0003 |
| TG(O-52:1) [NL-18:1] | DOWN | -42.98 | 63.20 | 101.09 | 0.0003 |
| PI 18:0_18:1 | DOWN | -28.03 | 1704.94 | 2382.75 | 0.0003 |
| SHexCer 18:1;O2/16:0 | DOWN | -27.97 | 2.34 | 3.21 | 0.0003 |
| CE 14:0 | DOWN | -25.23 | 6933.35 | 10097.24 | 0.0003 |
| LPC P-18:0 | DOWN | -22.07 | 132.79 | 167.27 | 0.0003 |
| TG(O-52:1) [NL-16:0] | DOWN | -42.44 | 79.11 | 119.19 | 0.0003 |
| SM 18:1;O2/20:0 \| SM 16:1;O2/22:0 | DOWN | -17.35 | 11594.33 | 14452.24 | 0.0003 |
| dimethyl-CE 20:4 | DOWN | -32.74 | 1533.66 | 2825.25 | 0.0003 |
| LPC 20:3 sn2 | DOWN | -24.58 | 615.77 | 724.84 | 0.0003 |
| PC P-35:2 b | DOWN | -22.15 | 142.06 | 180.50 | 0.0003 |
| HexCer 16:1;O2/22:0 | DOWN | -21.61 | 31.95 | 46.41 | 0.0003 |
| CE 18:2 | DOWN | -15.72 | 2207125.91 | 2561369.53 | 0.0003 |
| PE P-18:0/22:5 n3 | DOWN | -5.27 | 263.12 | 405.50 | 0.0003 |
| PC O-36:5 | DOWN | -26.73 | 712.13 | 1052.12 | 0.0004 |
| SM 18:1;O2/22:0 \| SM 16:1;O2/24:0 | DOWN | -18.59 | 18660.38 | 24053.07 | 0.0004 |
| CE 20:0 | DOWN | -25.64 | 225.43 | 311.47 | 0.0004 |
| PC 16:0_20:3 a | DOWN | -21.11 | 59376.39 | 77913.63 | 0.0004 |
| CE 20:3 | DOWN | -19.98 | 126889.21 | 170742.54 | 0.0004 |
| HexCer 18:2;O2/24:0 | DOWN | -20.90 | 88.18 | 125.41 | 0.0004 |
| methyl-CE 18:2 | DOWN | -30.85 | 12075.22 | 16740.74 | 0.0004 |
| PC O-16:0/20:3 | DOWN | -21.51 | 2509.19 | 3402.91 | 0.0004 |
| PE P-18:1/22:6 a | DOWN | -23.11 | 465.49 | 689.80 | 0.0004 |
| Ubiquinone-9 | DOWN | -35.38 | 116.77 | 210.47 | 0.0004 |
| methyl-DE 18:1 | DOWN | -26.46 | 609.13 | 869.86 | 0.0004 |
| TG(O-52:2) [NL-16:0] | DOWN | -38.57 | 108.06 | 163.56 | 0.0004 |
| PC O-34:2 | DOWN | -22.74 | 4754.84 | 6881.75 | 0.0004 |
| Cer 19:1;O2/24:0 | DOWN | -25.51 | 206.37 | 268.32 | 0.0004 |
| LPE 18:2 sn2 | DOWN | -27.76 | 462.72 | 615.84 | 0.0004 |
| PC 16:0_18:3 b | DOWN | -27.14 | 2226.58 | 3056.13 | 0.0004 |
| SM 18:2;O2/24:0 | DOWN | -16.91 | 8739.16 | 11813.09 | 0.0004 |
| SM 43:1;O2 | DOWN | -17.81 | 619.59 | 803.38 | 0.0004 |
| PC 17:0_18:2 | DOWN | -19.37 | 5621.55 | 7168.81 | 0.0004 |
| PE P-16:0/20:4 | DOWN | -12.20 | 1837.64 | 2866.36 | 0.0005 |
| PE P-16:0/22:6 | DOWN | -21.56 | 958.10 | 1256.65 | 0.0005 |
| PE P-16:0/18:1 | DOWN | 20.64 | 351.83 | 484.79 | 0.0006 |
| PC O-18:0/18:2 | DOWN | -15.35 | 872.68 | 1190.73 | 0.0006 |
| CE 20:4 | DOWN | -19.42 | 1003210.25 | 1386442.82 | 0.0006 |
| PE O-38:5 b | DOWN | -23.80 | 68.47 | 100.54 | 0.0006 |
| TG(48:3) [NL-18:3] | DOWN | -49.38 | 464.94 | 1110.43 | 0.0006 |
| methyl-DE 18:2 | DOWN | -29.40 | 1271.08 | 1961.25 | 0.0006 |
| LPE 18:2 sn1 | DOWN | -26.93 | 1135.82 | 1471.62 | 0.0006 |
| LPC 20:3 sn1 | DOWN | -24.36 | 1463.36 | 1909.18 | 0.0006 |
| PE P-17:0/20:4 a | DOWN | -20.85 | 72.59 | 103.18 | 0.0006 |
| PC 16:1_22:6 | DOWN | -23.48 | 317.71 | 421.52 | 0.0007 |
| PE P-18:0/20:3 a | DOWN | -18.82 | 150.95 | 220.73 | 0.0007 |
| PC 18:1_20:3 | DOWN | -19.17 | 7502.07 | 9564.89 | 0.0007 |
| PE P-18:1/22:4 b | DOWN | 1.60 | 128.71 | 169.73 | 0.0007 |
| PC 18:0_20:3 | DOWN | -21.46 | 16775.82 | 23566.73 | 0.0008 |
| PC 16:0_18:3 a | DOWN | -29.01 | 5432.53 | 7189.21 | 0.0008 |
| PE P-15:0/22:6 a | DOWN | -30.08 | 46.59 | 61.59 | 0.0008 |
| TG(O-50:1) [SIM] | DOWN | -32.29 | 82.67 | 110.88 | 0.0008 |
| PI 18:0_20:3 b | DOWN | -19.94 | 364.32 | 493.21 | 0.0008 |
| PE P-18:1/18:1 a | DOWN | -0.34 | 188.56 | 248.50 | 0.0008 |
| SM 18:0;O2/14:0 | DOWN | -20.05 | 328.07 | 435.32 | 0.0008 |
| PC 31:1 | DOWN | -33.63 | 12.67 | 17.77 | 0.0008 |
| CAR 24:0 | DOWN | -21.39 | 8.33 | 10.73 | 0.0008 |
| Cer 19:1;O2/23:0 | DOWN | -22.46 | 79.69 | 105.65 | 0.0008 |
| LPC 15:0 sn1 | DOWN | -19.15 | 751.67 | 909.93 | 0.0009 |
| LPC 18:3 sn2 a | DOWN | -31.02 | 71.97 | 92.51 | 0.0009 |
| LPC 20:3 (104) | DOWN | -24.40 | 1374.70 | 1737.15 | 0.0009 |
| LPC 26:0 sn2 | DOWN | -20.11 | 9.11 | 11.49 | 0.0009 |
| PC 16:0_18:2 | DOWN | -18.01 | 466700.80 | 523303.16 | 0.0010 |
| PC 17:1_18:2 | DOWN | -20.48 | 378.97 | 509.53 | 0.0010 |
| PC 31:0 a | DOWN | -33.65 | 248.67 | 352.86 | 0.0010 |
| TG(O-52:2) [NL-18:1] | DOWN | -34.89 | 110.85 | 166.70 | 0.0010 |
| PE O-16:0/20:3 | DOWN | -17.22 | 43.36 | 55.13 | 0.0010 |
| Cer 18:0;O/23:0 | DOWN | -24.86 | 101.73 | 150.50 | 0.0011 |
| PC O-34:4 | DOWN | -20.37 | 16.17 | 19.97 | 0.0011 |
| SM 18:2;O2/16:0 | DOWN | -12.29 | 17147.84 | 20712.02 | 0.0011 |
| PC 16:1_20:4 | DOWN | -18.45 | 823.15 | 1155.26 | 0.0011 |
| PC 15:0_22:6 | DOWN | -24.94 | 266.12 | 339.54 | 0.0012 |
| CE 22:0 | DOWN | -22.69 | 108.72 | 151.81 | 0.0012 |
| SPBP 16:1;O2 | DOWN | -18.66 | 56.91 | 73.81 | 0.0012 |
| PC 16:1_18:2 | DOWN | -22.21 | 4741.88 | 5976.37 | 0.0012 |
| LPC 20:0 sn1 | DOWN | -20.90 | 90.49 | 119.20 | 0.0013 |
| CE 16:2 | DOWN | -21.58 | 2581.23 | 3732.45 | 0.0013 |
| Cer 18:0;O/24:0 | DOWN | -18.50 | 240.13 | 322.83 | 0.0014 |
| PI 16:0_20:3 a | DOWN | -23.32 | 554.97 | 740.78 | 0.0014 |
| TG(48:2) [NL-18:2] | DOWN | -43.33 | 5810.44 | 9263.33 | 0.0015 |
| Cer 18:0;O2/22:0 | DOWN | -21.66 | 219.73 | 284.74 | 0.0016 |
| TG(O-54:2) [NL-18:1] | DOWN | -37.08 | 47.09 | 71.65 | 0.0017 |
| LPC 20:0 sn2 | DOWN | -19.52 | 24.69 | 31.64 | 0.0017 |
| SM 18:1;O2/24:0 | DOWN | -17.95 | 19334.51 | 25440.54 | 0.0017 |
| PE O-18:1/18:2 | DOWN | -18.38 | 70.51 | 99.17 | 0.0017 |
| PC 15-MHDA_20:4 | DOWN | -19.49 | 958.30 | 1241.80 | 0.0018 |
| Cer 17:1;O2/22:0 | DOWN | -19.27 | 79.15 | 103.45 | 0.0018 |
| LPC 17:1 sn1 a \| LPC 17:1 sn2 b | DOWN | -17.21 | 135.56 | 176.91 | 0.0018 |
| PC P-18:0/22:6 | DOWN | -23.71 | 188.42 | 242.87 | 0.0018 |
| TG(48:0) [NL-18:0] | DOWN | -37.47 | 751.74 | 1154.14 | 0.0018 |
| PC P-18:1/22:6 | DOWN | -23.43 | 127.34 | 166.24 | 0.0019 |
| CE 17:0 | DOWN | -21.81 | 6021.21 | 7113.54 | 0.0021 |
| PC 14:0_16:0 | DOWN | -22.85 | 3598.19 | 5041.79 | 0.0022 |
| PC 18:1_18:2 | DOWN | -22.55 | 35336.47 | 46023.24 | 0.0022 |
| CE 18:1 | DOWN | -13.78 | 439037.38 | 517573.40 | 0.0023 |
| PE P-17:0/20:4 b | DOWN | -10.77 | 98.45 | 135.02 | 0.0023 |
| methyl-CE 18:0 | DOWN | -17.09 | 295.48 | 345.15 | 0.0023 |
| PC P-18:0/20:4 | DOWN | -18.60 | 1521.21 | 1894.67 | 0.0024 |
| LPC 16:1 sn1 | DOWN | -16.41 | 1932.14 | 2508.20 | 0.0024 |
| SM 18:2;O2/18:1 | DOWN | -10.21 | 532.86 | 670.16 | 0.0024 |
| PC 15-MHDA_18:2 | DOWN | -21.24 | 2748.44 | 3648.52 | 0.0026 |
| PI 16:0_20:3 b | DOWN | -22.97 | 172.78 | 219.09 | 0.0026 |
| PE P-20:1/22:6 | DOWN | -18.56 | 32.93 | 42.84 | 0.0027 |
| PC P-36:3 | DOWN | -14.36 | 3251.48 | 3963.28 | 0.0027 |
| PE P-18:1/22:6 b | DOWN | -21.45 | 52.54 | 68.73 | 0.0028 |
| HexCer 18:1;O2/22:0 | DOWN | -14.94 | 325.31 | 403.67 | 0.0028 |
| SM 42:3;O2 | DOWN | -16.81 | 40857.25 | 49913.25 | 0.0029 |
| LPC P-16:0 | DOWN | -15.31 | 648.32 | 832.60 | 0.0029 |
| TG(50:4) [NL-20:4] | DOWN | -50.89 | 407.81 | 722.67 | 0.0030 |
| PC P-35:2 a | DOWN | -22.06 | 205.00 | 258.01 | 0.0031 |
| SM 18:2;O2/20:0 | DOWN | -11.39 | 4627.19 | 5625.88 | 0.0031 |
| PI 34:1 | DOWN | -25.03 | 1313.47 | 1759.59 | 0.0034 |
| PC 38:5 a | DOWN | -12.70 | 21940.10 | 26442.86 | 0.0034 |
| PC P-15:0/20:4 b | DOWN | -33.22 | 16.25 | 25.27 | 0.0034 |
| PE P-20:0/18:2 | DOWN | -12.45 | 136.98 | 184.91 | 0.0034 |
| TG(O-54:3) [NL-18:1] | DOWN | -27.69 | 43.30 | 59.36 | 0.0034 |
| HexCer 16:1;O2/20:0 | DOWN | -18.73 | 17.49 | 22.73 | 0.0035 |
| HexCer 18:1;O2/24:0 | DOWN | -17.19 | 410.02 | 528.12 | 0.0035 |
| Cer 18:1;O/23:0 | DOWN | -22.89 | 187.44 | 264.48 | 0.0036 |
| LPC 18:1 sn2 | DOWN | -17.41 | 5365.52 | 6501.82 | 0.0036 |
| PC 18:0_18:1 | DOWN | -13.47 | 18188.93 | 22974.71 | 0.0037 |
| TG(56:8) [NL-20:5] | DOWN | -11.12 | 921.22 | 1228.12 | 0.0037 |
| PE P-18:1/22:5 a | DOWN | -9.12 | 244.04 | 304.28 | 0.0037 |
| TG(48:3) [SIM] | DOWN | -39.60 | 5035.33 | 7596.39 | 0.0037 |
| CE 20:2 | DOWN | -15.72 | 2820.08 | 3609.44 | 0.0038 |
| LPC 18:1 sn1 | DOWN | -16.86 | 13697.68 | 17096.65 | 0.0039 |
| LPC P-17:0 a | DOWN | -17.71 | 29.01 | 34.68 | 0.0039 |
| LPC P-17:0 b | DOWN | -17.71 | 29.01 | 34.68 | 0.0039 |
| TG(48:3) [NL-14:0] | DOWN | -40.64 | 2066.63 | 2933.58 | 0.0039 |
| LPC 20:2 sn2 | DOWN | -17.50 | 66.79 | 77.64 | 0.0043 |
| LPC P-20:0 | DOWN | -14.90 | 7.71 | 9.68 | 0.0043 |
| PC 38:2 | DOWN | -13.29 | 2226.29 | 2716.90 | 0.0044 |
| PE P-20:1/20:4 | DOWN | -8.16 | 70.77 | 94.46 | 0.0044 |
| PC 18:0_20:4 | DOWN | -15.43 | 97004.80 | 110763.81 | 0.0045 |
| PI 18:0_22:4 | DOWN | -14.69 | 361.89 | 447.71 | 0.0045 |
| PI 36:2 | DOWN | -19.54 | 6511.77 | 8262.15 | 0.0045 |
| Cer 18:1;O/24:0 | DOWN | -23.61 | 581.89 | 796.60 | 0.0046 |
| PC O-18:1/18:2 | DOWN | -17.29 | 2779.25 | 3495.35 | 0.0046 |
| PE P-20:0/22:6 | DOWN | -18.39 | 92.26 | 114.23 | 0.0047 |
| PE P-15:0/22:6 b | DOWN | -20.22 | 30.11 | 35.66 | 0.0048 |
| PC P-38:5 a | DOWN | -11.77 | 3415.30 | 4127.35 | 0.0048 |
| SM 18:2;O2/17:0 | DOWN | -12.23 | 460.40 | 549.52 | 0.0049 |
| DG 14:0_18:2 | DOWN | -34.71 | 240.33 | 325.18 | 0.0051 |
| PC 18:0_22:5 n3 \| PC 20:1_20:4 | DOWN | -16.43 | 8068.93 | 10750.09 | 0.0051 |
| PC P-16:0/22:6 | DOWN | -20.07 | 894.52 | 1155.96 | 0.0051 |
| PI 15-MHDA_18:1 \| PI 17:0_18:1 | DOWN | -20.02 | 249.97 | 326.04 | 0.0051 |
| TG(48:1) [NL-18:1] | DOWN | -41.43 | 12826.81 | 19159.27 | 0.0051 |
| TG(52:5) [NL-20:5] | DOWN | -33.55 | 266.68 | 398.69 | 0.0053 |
| LPE P-18:0 | DOWN | 6.21 | 194.86 | 240.86 | 0.0055 |
| TG(50:4) [NL-14:0] | DOWN | -35.66 | 4195.50 | 6345.93 | 0.0058 |
| LPE 18:1 sn2 | DOWN | -20.83 | 247.90 | 304.09 | 0.0058 |
| Ubiquinone | DOWN | -25.21 | 2382.46 | 3190.67 | 0.0058 |
| LPC 17:1 sn2 | DOWN | -14.09 | 37.65 | 45.96 | 0.0059 |
| PE P-17:0/22:6 b | DOWN | -19.98 | 58.46 | 72.36 | 0.0060 |
| PE O-16:0/22:4 | DOWN | 2.84 | 67.03 | 81.95 | 0.0060 |
| SM 17:1;O2/16:0 | DOWN | -13.77 | 6339.39 | 7570.44 | 0.0061 |
| PC 17:0_20:4 | DOWN | -14.38 | 2027.31 | 2407.45 | 0.0066 |
| TG(50:4) [NL-18:3] | DOWN | -32.40 | 3162.19 | 4654.12 | 0.0066 |
| TG(O-54:4) [NL-18:2] | DOWN | -21.84 | 20.91 | 27.73 | 0.0070 |
| LPC 22:5 sn2 n3 | DOWN | -19.86 | 139.53 | 150.57 | 0.0070 |
| LPC 20:2 sn1 | DOWN | -18.07 | 176.25 | 219.44 | 0.0070 |
| SPBP 18:2;O2 | DOWN | -16.66 | 124.82 | 150.08 | 0.0070 |
| TG(48:2) [NL-14:0] | DOWN | -40.56 | 5764.35 | 8821.51 | 0.0071 |
| Cer 18:1;O2/26:0 | DOWN | -11.27 | 47.65 | 55.20 | 0.0075 |
| LPC P-18:1 | DOWN | -17.57 | 50.73 | 58.92 | 0.0075 |
| Cer 18:1;O2/22:0 | DOWN | -15.90 | 1536.56 | 1863.78 | 0.0075 |
| PC P-15:0/20:4 a | DOWN | -23.17 | 9.57 | 12.08 | 0.0075 |
| PC 38:4 b | DOWN | -16.57 | 6257.35 | 7941.13 | 0.0078 |
| Cer 16:1;O2/24:1 | DOWN | -14.55 | 133.36 | 161.86 | 0.0078 |
| PI 18:0_20:3 a | DOWN | -16.60 | 10308.05 | 11065.25 | 0.0078 |
| LPE P-16:0 | DOWN | 1.02 | 169.79 | 212.40 | 0.0078 |
| PC 39:5 a | DOWN | -24.33 | 31.22 | 39.80 | 0.0080 |
| CE 22:5 | DOWN | -13.79 | 16273.28 | 19988.79 | 0.0080 |
| LPE 18:0 sn1 | DOWN | -9.04 | 995.95 | 1261.59 | 0.0081 |
| LPC 16:1 sn2 | DOWN | -16.01 | 484.09 | 622.86 | 0.0081 |
| PC 38:5 b | DOWN | -13.68 | 6679.26 | 8725.33 | 0.0081 |
| TG(50:1) [NL-14:0] | DOWN | -41.23 | 920.88 | 1596.82 | 0.0083 |
| DG 18:1_20:5 | DOWN | -12.04 | 95.31 | 131.29 | 0.0085 |
| methyl-CE 20:4 | DOWN | -20.07 | 1384.88 | 2029.62 | 0.0086 |
| LPE 20:4 sn2 | DOWN | -13.98 | 360.98 | 427.66 | 0.0086 |
| TG(48:2) [SIM] | DOWN | -39.93 | 8784.79 | 11924.31 | 0.0086 |
| PC P-16:0/20:4 | DOWN | -11.39 | 9472.77 | 10734.56 | 0.0091 |
| SHexCer 18:1;O2/16:0;O | DOWN | -22.82 | 1.75 | 2.10 | 0.0092 |
| PC P-17:0/20:4 b | DOWN | -18.59 | 232.29 | 297.18 | 0.0093 |
| PE P-17:0/22:6 a | DOWN | -16.85 | 68.20 | 85.13 | 0.0093 |
| CE 15:0 | DOWN | -18.10 | 4205.13 | 5054.66 | 0.0095 |
| LPE 18:1 sn1 | DOWN | -19.35 | 843.01 | 1055.25 | 0.0096 |
| PC 15-MHDA_22:6 | DOWN | -24.33 | 232.05 | 296.84 | 0.0096 |
| LPC O-22:0 | DOWN | -10.92 | 75.34 | 92.58 | 0.0098 |
| TG(O-54:3) [NL-17:1] | DOWN | -30.38 | 15.02 | 19.19 | 0.0098 |
| SM 18:0;O2/22:0 | DOWN | -17.64 | 458.46 | 599.02 | 0.0099 |
| PC 16:0_18:0 | DOWN | -6.71 | 1941.86 | 2296.50 | 0.0100 |
| TG(54:6) [NL-20:5] | DOWN | -18.37 | 1270.37 | 1830.38 | 0.0107 |
| CE 22:6 | DOWN | -22.62 | 161035.23 | 198966.60 | 0.0110 |
| SM 18:0;O2/24:0 | DOWN | -17.62 | 89.99 | 108.61 | 0.0110 |
| TG(50:3) [NL-14:0] | DOWN | -31.55 | 10515.59 | 14681.25 | 0.0116 |
| PC 33:0 b | DOWN | -10.25 | 204.73 | 245.97 | 0.0118 |
| Cer 18:0;O/22:0 | DOWN | -12.72 | 257.31 | 316.77 | 0.0119 |
| TG(50:4) [SIM] | DOWN | -29.53 | 5891.27 | 8363.17 | 0.0120 |
| LPC 19:0 sn2 a | DOWN | -12.16 | 5.63 | 6.53 | 0.0124 |
| LPC 22:6 sn1 | DOWN | -22.08 | 799.54 | 945.99 | 0.0129 |
| SM 37:1;O2 | DOWN | -14.34 | 846.10 | 1035.26 | 0.0129 |
| LPC 20:4 sn1 | DOWN | -15.18 | 4772.60 | 5523.53 | 0.0131 |
| TG(O-52:2) [NL-17:1] | DOWN | -27.88 | 39.96 | 58.03 | 0.0133 |
| CE 20:1 | DOWN | -16.78 | 1290.79 | 1631.39 | 0.0134 |
| PI 18:0_22:5 n3 | DOWN | -16.08 | 683.25 | 822.55 | 0.0136 |
| CAR 18:0 | DOWN | -1.93 | 38.18 | 45.70 | 0.0136 |
| TG(54:7) [NL-20:5] | DOWN | -15.83 | 1278.22 | 1761.26 | 0.0136 |
| PE P-20:0/20:4 | DOWN | -0.50 | 349.86 | 423.80 | 0.0137 |
| TG(50:2) [NL-14:0] | DOWN | -27.32 | 12230.32 | 16974.48 | 0.0137 |
| LPC 22:5 n3 sn 1 (104) | DOWN | -18.89 | 268.71 | 330.66 | 0.0138 |
| LPC 22:5 sn1 n3 \| LPC 22:5 sn2 n6 | DOWN | -18.25 | 306.18 | 335.12 | 0.0141 |
| PC O-16:0/22:6 | DOWN | -17.30 | 2196.06 | 2527.39 | 0.0141 |
| SPBP 18:0;O2 | DOWN | -13.04 | 50.91 | 65.22 | 0.0141 |
| TG(50:3) [NL-18:3] | DOWN | -30.12 | 2052.50 | 3005.44 | 0.0141 |
| LPC O-18:0 | DOWN | -1.22 | 331.36 | 435.48 | 0.0142 |
| PC O-16:0/20:4 | DOWN | -9.96 | 20767.41 | 24707.64 | 0.0145 |
| PE P-18:1/22:4 a | DOWN | 13.00 | 134.82 | 165.05 | 0.0150 |
| Cer 16:1;O2/20:0 | DOWN | -12.03 | 30.90 | 36.64 | 0.0150 |
| Cer 19:1;O2/22:0 | DOWN | -15.62 | 93.02 | 114.51 | 0.0150 |
| PI 18:0_20:4 | DOWN | -10.12 | 24227.39 | 27273.25 | 0.0154 |
| TG(56:7) [NL-20:5] | DOWN | -9.20 | 1272.08 | 1582.63 | 0.0157 |
| Hex2Cer 16:1;O2/16:0 | DOWN | -12.70 | 36.09 | 44.54 | 0.0169 |
| PI 18:1_18:2 | DOWN | -14.06 | 438.71 | 585.24 | 0.0178 |
| PI 16:0/16:0 | DOWN | -22.94 | 111.46 | 126.02 | 0.0180 |
| Cer 17:1;O2/24:1 | DOWN | -12.62 | 122.90 | 148.30 | 0.0181 |
| LPC 22:6 sn2 | DOWN | -19.65 | 402.31 | 446.09 | 0.0188 |
| Cer 20:1;O2/24:0 | DOWN | -16.54 | 38.32 | 45.60 | 0.0189 |
| PC 17:0_22:6 | DOWN | -19.61 | 360.85 | 432.60 | 0.0189 |
| TG(48:1) [SIM] | DOWN | -40.42 | 5221.58 | 7316.95 | 0.0191 |
| PC O-18:0/20:4 | DOWN | -9.08 | 3896.68 | 4519.49 | 0.0202 |
| LPC O-16:0 | DOWN | -4.98 | 1327.59 | 1648.41 | 0.0203 |
| LPC O-20:0 | DOWN | -3.40 | 71.18 | 89.82 | 0.0206 |
| TG(48:2) [NL-14:1] | DOWN | -40.99 | 1876.57 | 2719.40 | 0.0206 |
| CE 17:1 | DOWN | -16.04 | 13093.50 | 15502.46 | 0.0213 |
| PC O-18:0/22:6 | DOWN | -16.34 | 954.81 | 1073.34 | 0.0220 |
| SM 40:3;O2 b | DOWN | -9.13 | 692.01 | 835.86 | 0.0220 |
| PE P-16:0/22:4 | DOWN | 22.39 | 400.38 | 505.20 | 0.0222 |
| DE 18:1 | DOWN | -3.67 | 979.06 | 1167.30 | 0.0222 |
| TG(48:3) [NL-16:1] | DOWN | -30.19 | 1761.47 | 2457.67 | 0.0222 |
| DG 14:0_16:0 | DOWN | -30.26 | 159.48 | 197.44 | 0.0231 |
| PC 16:0_22:6 | DOWN | -17.01 | 33467.07 | 38327.33 | 0.0237 |
| SM 18:0;O2/24:1 | DOWN | -14.00 | 3121.09 | 3775.69 | 0.0237 |
| PI 15-MHDA_18:2 \| PI 17:0_18:2 | DOWN | -13.34 | 180.82 | 203.17 | 0.0246 |
| PC 16:0_20:4 | DOWN | -9.22 | 197154.27 | 217104.95 | 0.0256 |
| PC 17:0_18:1 | DOWN | -10.15 | 2290.85 | 2755.69 | 0.0256 |
| PE P-18:0/22:4 | DOWN | 40.71 | 167.68 | 209.78 | 0.0257 |
| TG(56:9) [SIM] | DOWN | -13.39 | 477.31 | 610.98 | 0.0259 |
| LPC 20:4 sn2 | DOWN | -13.77 | 2206.22 | 2488.25 | 0.0277 |
| LPI 18:2 sn1 | DOWN | -21.69 | 25.27 | 27.23 | 0.0285 |
| PC P-16:0/18:0 | DOWN | -5.90 | 47.57 | 52.04 | 0.0285 |
| TG(49:1) [SIM] | DOWN | -35.89 | 3552.90 | 4451.45 | 0.0285 |
| TG(50:3) [NL-14:1] | DOWN | -27.82 | 1390.25 | 1780.99 | 0.0285 |
| PE 16:0_18:3 b | DOWN | -21.48 | 44.95 | 57.42 | 0.0294 |
| PC P-38:5 b | DOWN | -15.49 | 410.60 | 489.46 | 0.0295 |
| TG(58:10) [SIM] | DOWN | -8.28 | 264.26 | 337.72 | 0.0297 |
| TG(48:0) [SIM] | DOWN | -22.12 | 1696.83 | 2080.75 | 0.0298 |
| PI 18:0_20:2 | DOWN | -11.78 | 240.44 | 279.91 | 0.0307 |
| CE 18:2 [+OH] | DOWN | -18.28 | 5351.46 | 7428.16 | 0.0311 |
| PC 33:1 | DOWN | -14.16 | 2798.50 | 3422.56 | 0.0315 |
| LPE 16:0 sn1 | DOWN | -13.07 | 767.85 | 876.74 | 0.0329 |
| Cer 18:2;O2/24:1 | DOWN | -6.40 | 263.99 | 324.32 | 0.0335 |
| PC 20:0_20:4 | DOWN | -3.69 | 713.68 | 798.72 | 0.0335 |
| PI 16:0_20:4 | DOWN | -12.95 | 2012.19 | 2347.15 | 0.0345 |
| PI 15-MHDA_20:4 \| PI 17:0_20:4 | DOWN | -13.09 | 258.34 | 318.30 | 0.0349 |
| Cer 18:1;O/22:0 | DOWN | -19.17 | 447.64 | 568.50 | 0.0356 |
| LPC O-24:0 | DOWN | -8.86 | 291.00 | 327.21 | 0.0358 |
| PC P-17:0/20:4 a | DOWN | -14.16 | 502.49 | 590.01 | 0.0362 |
| PE 15-MHDA_18:2 | DOWN | -18.81 | 30.78 | 38.59 | 0.0362 |
| PC 18:1_22:6 b | DOWN | -15.47 | 488.46 | 566.37 | 0.0387 |
| PC 38:7 c | DOWN | -13.37 | 107.41 | 131.05 | 0.0387 |
| CE 24:0 | DOWN | -8.99 | 86.62 | 99.99 | 0.0413 |
| PC 18:0_22:4 | DOWN | -11.54 | 4294.45 | 5131.60 | 0.0434 |
| LPE 20:4 sn1 | DOWN | -9.00 | 659.48 | 763.22 | 0.0435 |
| PC 18:1_22:6 a | DOWN | -16.38 | 2895.74 | 3140.30 | 0.0435 |
| CE 22:4 | DOWN | -9.06 | 5201.08 | 6779.63 | 0.0441 |
| PE 16:0_20:5 | DOWN | -10.21 | 94.81 | 110.05 | 0.0452 |
| TG(52:5) [NL-18:3] | DOWN | -23.60 | 11531.68 | 15153.16 | 0.0452 |
| PI 38:5 b | DOWN | -13.48 | 654.26 | 771.36 | 0.0454 |
| LPC 19:1 b | DOWN | -10.40 | 51.94 | 61.49 | 0.0460 |
| TG(O-52:2) [SIM] | DOWN | -25.39 | 66.47 | 80.34 | 0.0465 |
| PC 18:0_22:6 | DOWN | -16.09 | 9800.29 | 11558.45 | 0.0474 |
| SM 18:0;O2/20:0 | DOWN | -13.34 | 448.57 | 555.57 | 0.0480 |
| Hex2Cer 18:2;O2/16:0 | DOWN | -11.39 | 45.65 | 53.19 | 0.0482 |
| SPBP 18:1;O2 | DOWN | -10.80 | 407.55 | 500.09 | 0.0496 |
| dxCA | DOWN | -21.76 | 289.33 | 386.61 | 0.0558 |
| PC P-18:0/22:5 | DOWN | -11.57 | 656.11 | 728.94 | 0.0558 |
| LPC 22:4 sn2 | DOWN | -11.53 | 81.86 | 90.05 | 0.0572 |
| TG(52:4) [NL-18:3] | DOWN | -20.26 | 14750.95 | 17881.06 | 0.0583 |
| DG 18:1_20:3 | DOWN | -17.47 | 589.55 | 763.05 | 0.0601 |
| PI 38:5 a | DOWN | -8.38 | 1164.47 | 1343.85 | 0.0607 |
| SM 18:0;O2/16:0 | DOWN | -9.54 | 6194.27 | 6975.38 | 0.0607 |
| PC 18:1/18:1 | DOWN | -8.51 | 15145.83 | 17298.61 | 0.0614 |
| LPE 22:6 sn2 | DOWN | -9.54 | 450.56 | 520.44 | 0.0626 |
| SM 37:2;O2 | DOWN | -8.59 | 191.90 | 223.71 | 0.0675 |
| SM 16:1;O2/24:1 | DOWN | -8.83 | 5077.60 | 5739.46 | 0.0678 |
| TG(O-54:4) [NL-17:1] | DOWN | -20.40 | 12.63 | 15.93 | 0.0710 |
| TG(50:3) [NL-18:2] | DOWN | -22.61 | 26097.61 | 31488.45 | 0.0718 |
| Hex2Cer 18:1;O2/24:1 | DOWN | 3.47 | 98.47 | 111.23 | 0.0727 |
| PC 15-MHDA_18:1 | DOWN | -12.35 | 605.83 | 705.88 | 0.0733 |
| CE 22:1 | DOWN | -10.61 | 258.87 | 324.11 | 0.0734 |
| TG(50:0) [SIM] | DOWN | -12.68 | 1192.69 | 1400.34 | 0.0734 |
| PI 18:0_22:6 | DOWN | -11.23 | 886.40 | 1010.43 | 0.0746 |
| TG(52:5) [SIM] | DOWN | -20.74 | 8110.84 | 10498.74 | 0.0746 |
| PC 31:0 b | DOWN | -9.83 | 632.52 | 732.74 | 0.0763 |
| PC 34:2 [+OH] | DOWN | -14.44 | 19.24 | 25.06 | 0.0793 |
| TG(50:3) [SIM] | DOWN | -21.44 | 14423.22 | 16676.62 | 0.0829 |
| PC P-16:0/14:0 | DOWN | -5.35 | 120.67 | 135.80 | 0.0844 |
| PI 16:0_16:1 | DOWN | -23.79 | 210.95 | 268.11 | 0.0847 |
| PE P-18:0/22:5 n6 | DOWN | -8.37 | 98.32 | 114.05 | 0.0857 |
| LPE 16:0 sn2 | DOWN | -9.53 | 293.24 | 322.53 | 0.0860 |
| LPE P-18:1 | DOWN | -0.82 | 72.09 | 85.98 | 0.0876 |
| PC P-18:1/18:1 | DOWN | -9.77 | 410.34 | 441.71 | 0.0903 |
| LPE P-20:0 | DOWN | 6.44 | 22.32 | 26.17 | 0.0919 |
| PE P-18:1/20:5 b | DOWN | -18.07 | 18.10 | 20.68 | 0.1015 |
| CAR 15:0 a | DOWN | -12.39 | 1.63 | 1.83 | 0.1020 |
| CE 24:5 | DOWN | 8.21 | 465.31 | 599.60 | 0.1020 |
| Cer 16:1;O2/16:0 | DOWN | -4.45 | 19.80 | 23.24 | 0.1020 |
| LPE 22:6 sn1 | DOWN | -10.97 | 494.24 | 552.19 | 0.1020 |
| PE P-16:0/22:5 n6 | DOWN | -8.27 | 281.71 | 312.69 | 0.1020 |
| TG(52:5) [NL-20:4] | DOWN | -30.85 | 1807.86 | 2098.79 | 0.1020 |
| CE 24:6 | DOWN | -11.34 | 353.44 | 411.02 | 0.1049 |
| PC 33:0 a | DOWN | -13.13 | 456.58 | 542.72 | 0.1049 |
| PC O-36:0 | DOWN | 17.57 | 43.49 | 51.77 | 0.1074 |
| TG(49:1) [NL-16:1] | DOWN | -33.61 | 245.98 | 273.74 | 0.1078 |
| TG(51:2) [NL-17:0] | DOWN | -25.67 | 1461.83 | 1730.26 | 0.1082 |
| Cer 20:1;O2/23:0 | DOWN | -12.32 | 13.82 | 13.93 | 0.1126 |
| PC 16:0_18:1 | DOWN | -8.24 | 196683.06 | 215736.73 | 0.1131 |
| PE 18:0_18:2 | DOWN | -19.36 | 3852.49 | 4478.21 | 0.1143 |
| CE 16:0 | DOWN | -5.06 | 59326.25 | 61333.92 | 0.1184 |
| LPE 18:0 sn2 | DOWN | -3.48 | 433.59 | 486.77 | 0.1186 |
| TG(51:2) [NL-15:0] | DOWN | -20.50 | 3580.60 | 3863.26 | 0.1186 |
| Hex2Cer 18:1;O2/24:0 | DOWN | 17.44 | 172.91 | 202.63 | 0.1243 |
| CE 20:4 [+OH] | DOWN | -6.18 | 907.36 | 1184.43 | 0.1269 |
| TG(56:8) [SIM] | DOWN | -10.52 | 1436.89 | 1668.75 | 0.1269 |
| CAR 26:0 | DOWN | -12.02 | 15.51 | 17.05 | 0.1279 |
| TG(52:2) [NL-18:2] | DOWN | -20.27 | 5901.18 | 7715.83 | 0.1279 |
| TG(48:2) [NL-16:1] | DOWN | -28.71 | 5570.56 | 7483.60 | 0.1299 |
| TG(54:4) [NL-20:3] | DOWN | -19.59 | 3917.37 | 4109.93 | 0.1304 |
| PE 16:0_20:3 | DOWN | -12.70 | 346.98 | 421.32 | 0.1326 |
| LPI 18:2 sn2 | DOWN | -10.45 | 16.22 | 17.45 | 0.1372 |
| SM 41:1;O2 a | DOWN | -1.54 | 263.76 | 308.18 | 0.1377 |
| PE 16:1_20:4 | DOWN | -7.54 | 33.01 | 37.23 | 0.1382 |
| Cer 18:2;O2/20:0 | DOWN | -7.65 | 15.00 | 16.41 | 0.1427 |
| LPC 18:2 [+OH] | DOWN | -11.40 | 9.54 | 11.39 | 0.1427 |
| CAR 15:0 b | DOWN | -4.68 | 1.71 | 1.86 | 0.1446 |
| PC P-16:0/18:1 | DOWN | -6.89 | 2475.44 | 2768.69 | 0.1446 |
| TG(O-50:2) [NL-18:1] | DOWN | -29.35 | 22.15 | 25.57 | 0.1460 |
| LPC 22:4 sn1 | DOWN | -9.51 | 135.87 | 143.13 | 0.1475 |
| SM 44:1;O2 | DOWN | -2.97 | 1070.89 | 1237.27 | 0.1480 |
| TG(50:2) [NL-18:2] | DOWN | -18.80 | 23474.30 | 27156.16 | 0.1518 |
| Hex2Cer 18:2;O2/24:1 | DOWN | 0.57 | 40.12 | 44.88 | 0.1533 |
| LPC 20:1 sn1 | DOWN | -5.84 | 162.07 | 176.69 | 0.1535 |
| Cer 17:1;O2/16:0 | DOWN | -2.86 | 14.93 | 15.78 | 0.1547 |
| PE 15-MHDA_22:6 | DOWN | -14.38 | 29.17 | 31.76 | 0.1547 |
| LPC 22:5 sn1 n6 | DOWN | -9.03 | 96.71 | 101.88 | 0.1553 |
| TG(49:1) [NL-17:1] | DOWN | -28.76 | 923.55 | 1033.08 | 0.1558 |
| SM 38:3;O2 a | DOWN | -0.68 | 279.64 | 320.09 | 0.1564 |
| CAR 17:0 b | DOWN | 1.84 | 3.17 | 3.31 | 0.1570 |
| TG(48:1) [NL-16:1] | DOWN | -31.80 | 4962.02 | 6266.31 | 0.1594 |
| TG(51:2) [SIM] | DOWN | -20.43 | 8144.37 | 9303.72 | 0.1619 |
| TG(51:1) [SIM] | DOWN | -27.30 | 2804.88 | 3193.61 | 0.1692 |
| Cer 18:0;O/20:0 | DOWN | -3.56 | 98.59 | 112.83 | 0.1694 |
| LPI 20:4 sn1 | DOWN | -10.40 | 35.92 | 40.31 | 0.1694 |
| TG(52:1) [NL-18:0] | DOWN | -23.96 | 14039.54 | 17131.66 | 0.1701 |
| TG(54:7) [SIM] | DOWN | -21.92 | 2514.99 | 3063.27 | 0.1717 |
| PE 18:1_18:2 | DOWN | -14.16 | 862.25 | 1067.61 | 0.1723 |
| Hex3Cer 18:1;O2/22:0 | DOWN | -8.21 | 11.81 | 13.42 | 0.1729 |
| TG(56:8) [NL-20:4] | DOWN | -15.31 | 1638.88 | 1743.89 | 0.1745 |
| TG(54:7) [NL-22:6] | DOWN | -20.55 | 713.09 | 836.38 | 0.1788 |
| Hex2Cer 18:1;O2/16:0 | DOWN | 6.20 | 547.51 | 613.75 | 0.1866 |
| Cer 20:1;O2/22:0 | DOWN | -12.09 | 20.69 | 23.20 | 0.1969 |
| CE 16:1 | DOWN | -5.62 | 67486.24 | 74545.14 | 0.1993 |
| TG(54:1) [NL-18:1] | DOWN | -25.14 | 2182.07 | 2923.64 | 0.1993 |
| HexCer 18:2;O2/20:0 | DOWN | -5.04 | 17.90 | 20.20 | 0.2000 |
| PE 18:0_18:1 | DOWN | 2.31 | 775.66 | 780.75 | 0.2040 |
| PC O-40:7 | DOWN | -8.08 | 709.54 | 745.33 | 0.2047 |
| Cer 18:0;O/24:1 | DOWN | 4.86 | 196.32 | 214.12 | 0.2054 |
| TG(52:1) [SIM] | DOWN | -22.53 | 6248.71 | 7867.30 | 0.2080 |
| TG(56:7) [SIM] | DOWN | -7.17 | 2031.96 | 2272.79 | 0.2080 |
| TG(56:9) [NL-22:6] | DOWN | -12.82 | 478.68 | 559.48 | 0.2087 |
| TG(48:0) [NL-16:0] | DOWN | -12.55 | 11145.43 | 12675.21 | 0.2128 |
| TG(54:1) [SIM] | DOWN | -25.44 | 1045.06 | 1263.68 | 0.2135 |
| CE 24:1 | DOWN | -6.19 | 216.13 | 260.13 | 0.2143 |
| PC 32:1 | DOWN | -13.51 | 13162.96 | 14652.53 | 0.2196 |
| LPC 22:1 sn1 | DOWN | -11.72 | 16.50 | 17.54 | 0.2239 |
| Cer 16:1;O2/18:0 | DOWN | -7.68 | 45.39 | 53.92 | 0.2271 |
| TG(51:1) [NL-17:0] | DOWN | -23.70 | 1821.04 | 1993.92 | 0.2290 |
| PE 18:0_20:3 a | DOWN | -7.66 | 513.13 | 546.77 | 0.2511 |
| TG(50:2) [NL-18:1] | DOWN | -16.75 | 55078.50 | 61800.52 | 0.2545 |
| Cer 17:1;O2/20:0 | DOWN | -12.65 | 10.66 | 11.72 | 0.2553 |
| LPC 20:1 sn2 | DOWN | -5.90 | 47.50 | 49.82 | 0.2587 |
| DG 18:1_18:3 | DOWN | -13.61 | 343.99 | 381.72 | 0.2590 |
| TG(50:0) [NL-18:0] | DOWN | -10.93 | 2697.26 | 2949.34 | 0.2590 |
| SM 18:2;O2/18:0 | DOWN | -0.57 | 10151.16 | 10937.26 | 0.2665 |
| PI 38:6 | DOWN | -5.49 | 275.02 | 294.86 | 0.2701 |
| LPC 19:1 a | DOWN | -7.84 | 15.12 | 15.64 | 0.2736 |
| TG(54:5) [NL-18:3] | DOWN | -17.40 | 9653.38 | 10914.25 | 0.2772 |
| cholic acid | DOWN | -36.36 | 70.80 | 85.25 | 0.2781 |
| TG(52:1) [NL-18:1] | DOWN | -17.78 | 18065.78 | 22078.37 | 0.2812 |
| TG(52:4) [NL-18:2] | DOWN | -12.40 | 89569.61 | 98762.99 | 0.2812 |
| TG(54:6) [NL-18:3] | DOWN | -20.60 | 6966.34 | 7502.62 | 0.2820 |
| DG 16:0_18:2 | DOWN | -11.31 | 4082.28 | 4561.70 | 0.2829 |
| LPC O-24:2 | DOWN | -3.35 | 39.86 | 42.01 | 0.2837 |
| SM 18:1;O2/16:0 | DOWN | -3.66 | 104780.18 | 116338.40 | 0.2912 |
| TG(50:1) [SIM] | DOWN | -21.26 | 15543.45 | 17168.31 | 0.2912 |
| PG 36:2 | DOWN | -0.99 | 186.80 | 195.35 | 0.3111 |
| LPC O-20:1 | DOWN | 3.11 | 53.50 | 53.80 | 0.3180 |
| PC O-18:0/18:1 | DOWN | 19.73 | 346.70 | 385.47 | 0.3313 |
| SM 17:1;O2/24:1 | DOWN | -4.63 | 3550.16 | 3841.86 | 0.3338 |
| CAR 18:3 | DOWN | -4.13 | 3.39 | 3.86 | 0.3347 |
| DG 18:2_20:4 | DOWN | -12.07 | 454.72 | 461.84 | 0.3372 |
| HexCer 16:1;O2/18:0 | DOWN | -4.14 | 19.03 | 20.61 | 0.3455 |
| TG(50:2) [SIM] | DOWN | -15.37 | 14510.22 | 15455.60 | 0.3546 |
| TG(52:4) [SIM] | DOWN | -11.55 | 23363.43 | 26013.87 | 0.3605 |
| PC P-20:0/20:4 | DOWN | -4.61 | 82.15 | 87.81 | 0.3681 |
| DG 16:0_22:6 | DOWN | -8.75 | 85.40 | 100.55 | 0.3708 |
| FA 18:3 | DOWN | -9.96 | 5691.61 | 6052.81 | 0.3802 |
| DG 16:0_16:1 | DOWN | -20.46 | 314.60 | 333.06 | 0.3805 |
| TG(54:6) [NL-22:6] | DOWN | -21.51 | 664.34 | 826.73 | 0.3805 |
| DE 16:0 | DOWN | -2.88 | 285.37 | 308.77 | 0.3922 |
| TG(51:2) [NL-17:1] | DOWN | -16.74 | 5013.06 | 5336.57 | 0.3925 |
| PE P-18:1/22:5 b | DOWN | -1.28 | 70.68 | 71.82 | 0.3952 |
| DG 16:1_18:1 | DOWN | -10.43 | 716.81 | 811.66 | 0.3963 |
| TG(54:6) [SIM] | DOWN | -18.58 | 6215.86 | 6267.11 | 0.4043 |
| PE 17:0_18:1 | DOWN | -2.94 | 54.33 | 55.96 | 0.4089 |
| TG(53:2) [NL-18:1] | DOWN | -10.28 | 11046.55 | 11639.83 | 0.4092 |
| TG(54:2) [NL-20:1] | DOWN | -18.19 | 2556.78 | 3062.70 | 0.4092 |
| PE 16:1_18:2 | DOWN | -17.04 | 35.52 | 35.65 | 0.4166 |
| CAR 17:0 a | DOWN | -1.03 | 3.58 | 3.84 | 0.4213 |
| CAR 20:5 | DOWN | 1.20 | 0.48 | 0.56 | 0.4223 |
| TG(50:3) [NL-16:1] | DOWN | -14.70 | 21064.98 | 22746.51 | 0.4266 |
| Cer 18:2;O2/16:0 | DOWN | 4.26 | 102.89 | 111.68 | 0.4276 |
| PC 18:0_22:5 n6 | DOWN | -5.90 | 1254.01 | 1411.86 | 0.4287 |
| PE 17:0_18:2 | DOWN | -8.49 | 93.21 | 101.92 | 0.4363 |
| PIP 38:4 | DOWN | 4.41 | 78.83 | 82.95 | 0.4363 |
| DG 18:0_18:2 | DOWN | -10.46 | 1172.86 | 1238.08 | 0.4450 |
| LPC O-24:1 | DOWN | -3.85 | 152.18 | 153.73 | 0.4550 |
| DG 16:0_18:1 | DOWN | -8.65 | 6491.24 | 7565.68 | 0.4571 |
| PE 18:1_22:6 a | DOWN | -4.35 | 281.17 | 304.27 | 0.4571 |
| TG(54:2) [SIM] | DOWN | -15.95 | 6674.55 | 7053.47 | 0.4601 |
| PE 20:0_20:4 | DOWN | 20.14 | 23.90 | 25.90 | 0.4631 |
| Hex2Cer 18:1;O2/22:0 | DOWN | 19.80 | 39.72 | 42.65 | 0.4853 |
| PC O-18:1/18:1 | DOWN | 4.06 | 699.19 | 721.61 | 0.4957 |
| TG(52:4) [NL-16:1] | DOWN | -8.15 | 27135.42 | 28957.41 | 0.4957 |
| TG(54:5) [NL-20:4] | DOWN | -12.15 | 10920.57 | 11480.71 | 0.4957 |
| Cer 18:1;O2/21:0 | DOWN | 0.95 | 49.58 | 52.94 | 0.5158 |
| PC O-38:5 | DOWN | 0.02 | 10835.60 | 11183.29 | 0.5158 |
| SM 16:1;O2/19:0 | DOWN | -1.65 | 681.92 | 760.79 | 0.5170 |
| PE 38:5 b | DOWN | -7.24 | 602.19 | 653.98 | 0.5221 |
| CAR 13:0 | DOWN | -8.28 | 6.43 | 6.75 | 0.5235 |
| TG(50:1) [NL-18:1] | DOWN | -13.99 | 52714.99 | 54951.41 | 0.5235 |
| PE 17:0_22:6 | DOWN | -3.80 | 54.62 | 60.05 | 0.5287 |
| TG(52:3) [NL-18:2] | DOWN | -7.16 | 161367.09 | 165969.28 | 0.5319 |
| PE 38:5 a | DOWN | 5.02 | 1497.95 | 1577.97 | 0.5403 |
| PC P-16:0/16:1 | DOWN | 1.97 | 365.85 | 384.77 | 0.5497 |
| DG 16:0_20:4 | DOWN | -11.61 | 277.48 | 289.46 | 0.5508 |
| HexCer 18:2;O2/22:0 | DOWN | -2.91 | 42.89 | 47.28 | 0.5531 |
| PC O-32:1 | DOWN | 0.64 | 133.93 | 137.66 | 0.5531 |
| CE 24:4 | DOWN | 8.46 | 1426.59 | 1519.89 | 0.5542 |
| TG(53:2) [SIM] | DOWN | -10.37 | 3461.89 | 3704.88 | 0.5575 |
| Hex3Cer 18:1;O2/18:0 | DOWN | -3.19 | 30.34 | 30.69 | 0.5586 |
| SM 44:2;O2 | DOWN | 5.23 | 244.35 | 256.59 | 0.5597 |
| TG(54:4) [SIM] | DOWN | -9.05 | 6129.55 | 6458.66 | 0.5630 |
| CAR 20:4 | DOWN | 0.85 | 2.67 | 2.98 | 0.5835 |
| TG(54:5) [SIM] | DOWN | -12.00 | 7465.93 | 7614.12 | 0.5846 |
| DG 18:1_18:2 | DOWN | -6.99 | 11508.31 | 12486.56 | 0.5997 |
| PS 40:5 | DOWN | 139.34 | 89.50 | 97.18 | 0.5997 |
| DG 18:1_22:6 | DOWN | 1.27 | 163.02 | 166.02 | 0.6009 |
| Hex2NeuAcCer 18:1;O2/22:0 | DOWN | -0.88 | 34.89 | 35.55 | 0.6020 |
| Hex2Cer 16:1;O2/24:1 | DOWN | 2.41 | 14.26 | 15.04 | 0.6075 |
| DG 18:0_18:1 | DOWN | -7.47 | 1320.54 | 1407.09 | 0.6173 |
| CAR 20:3 | DOWN | 1.03 | 2.93 | 3.07 | 0.6295 |
| TG(50:1) [NL-16:0] | DOWN | -15.46 | 89739.59 | 90601.37 | 0.6295 |
| Cer 18:1;O/20:0 | DOWN | -5.14 | 234.01 | 251.85 | 0.6318 |
| TG(50:2) [NL-16:1] | DOWN | -11.84 | 30132.41 | 31282.18 | 0.6475 |
| TG(52:2) [NL-16:0] | DOWN | -4.92 | 185037.37 | 197874.73 | 0.6508 |
| TG(54:2) [NL-18:0] | DOWN | -10.00 | 30962.49 | 33335.06 | 0.6631 |
| TG(56:6) [SIM] | DOWN | 0.66 | 1578.70 | 1632.42 | 0.7004 |
| Cer 18:2;O2/18:0 | DOWN | 5.62 | 33.04 | 36.99 | 0.7152 |
| PE 16:0_16:1 | DOWN | -13.72 | 160.91 | 160.92 | 0.7198 |
| PE 18:0_20:3 b | DOWN | 11.30 | 42.21 | 44.17 | 0.7302 |
| PS 38:4 | DOWN | 129.90 | 376.79 | 429.69 | 0.7359 |
| TG(56:7) [NL-22:5] | DOWN | -4.16 | 2372.21 | 2465.69 | 0.7463 |
| CAR 10:0 | DOWN | -14.78 | 140.56 | 161.63 | 0.7474 |
| SM 43:2;O2 b | DOWN | 1.37 | 1950.04 | 1970.72 | 0.7509 |
| CAR 18:0;O | DOWN | 5.12 | 2.32 | 2.35 | 0.7531 |
| SM 19:1;O2/24:1 | DOWN | 1.47 | 1948.01 | 1992.05 | 0.7531 |
| PE 16:0_18:3 a | DOWN | -10.39 | 102.67 | 105.93 | 0.7612 |
| SM 40:3;O2 a | DOWN | 3.39 | 1263.68 | 1271.62 | 0.7657 |
| TG(54:3) [NL-18:2] | DOWN | -7.34 | 21427.73 | 22346.27 | 0.7691 |
| TG(56:6) [NL-20:4] | DOWN | 0.08 | 8146.19 | 8215.64 | 0.7725 |
| CAR 26:1 | DOWN | 1.01 | 4.89 | 5.04 | 0.7736 |
| PE 18:0_20:4 | DOWN | 9.31 | 7061.06 | 7235.42 | 0.7770 |
| CAR 16:0 | DOWN | 5.15 | 94.37 | 98.65 | 0.7805 |
| CAR 18:2 | DOWN | 4.13 | 57.86 | 58.71 | 0.7943 |
| TG(54:3) [SIM] | DOWN | -7.26 | 11710.82 | 12629.85 | 0.7954 |
| Hex2NeuAcCer 18:1;O2/20:0 | DOWN | 4.69 | 14.14 | 14.15 | 0.8128 |
| DG 18:1_18:1 | DOWN | 2.96 | 24964.77 | 27017.97 | 0.8173 |
| TG(56:6) [NL-22:5] | DOWN | -4.02 | 3795.36 | 4037.26 | 0.8265 |
| Cer 18:0;O2/16:0 | DOWN | -2.35 | 73.45 | 74.63 | 0.8401 |
| PE 18:1_22:6 b | DOWN | 1.07 | 105.82 | 112.02 | 0.8401 |
| TG(52:2) [SIM] | DOWN | -3.59 | 41193.57 | 44235.45 | 0.8401 |
| SM 18:1;O2/24:1 | DOWN | 4.76 | 33405.53 | 34135.59 | 0.8410 |
| DG 18:2_18:2 | DOWN | -11.26 | 1895.20 | 1902.51 | 0.8501 |
| HexCer 18:1;O2/18:0 | DOWN | 9.37 | 63.47 | 64.20 | 0.8626 |
| PS 36:1 | DOWN | 133.32 | 261.50 | 264.16 | 0.8626 |
| PC O-16:0/16:0 | DOWN | 13.11 | 2551.88 | 2610.44 | 0.8667 |
| Cer 19:1;O2/24:1 | DOWN | 5.88 | 166.84 | 170.94 | 0.9287 |
| DG 18:2_22:6 | DOWN | 1.56 | 89.48 | 94.63 | 0.9287 |
| Cer 18:1;O2/24:1 | DOWN | 8.75 | 1546.12 | 1576.18 | 0.9386 |
| PE 16:0_22:6 | DOWN | 1.03 | 3193.18 | 3384.29 | 0.9428 |
| PE 16:0_18:1 | DOWN | 8.61 | 2052.59 | 2098.39 | 0.9596 |
| PC 38:6 [+OH] | DOWN | -3.93 | 34.88 | 35.12 | 0.9614 |
| CAR 16:0;O | DOWN | 4.64 | 2.81 | 2.83 | 0.9633 |
| COH | DOWN | 0.41 | 146185.83 | 146256.41 | 0.9633 |
| PC O-34:1 | DOWN | 8.99 | 4117.34 | 4184.02 | 0.9651 |
| PC 16:0_16:0 | DOWN | 5.47 | 15033.96 | 15276.84 | 0.9660 |
| PE 18:0_22:6 | DOWN | -1.33 | 1832.85 | 1874.24 | 0.9669 |
| FA 18:2 | DOWN | -4.97 | 106128.24 | 107361.00 | 0.9926 |
| Hex2Cer 18:1;O2/20:0 | UP | 23.50 | 11.90 | 10.01 | 0.0204 |
| CAR 14:1;O | UP | 14.07 | 7.62 | 6.07 | 0.0251 |
| Cer 19:1;O2/18:0 | UP | 27.07 | 5.91 | 4.72 | 0.0369 |
| Cer 18:1;O2/18:0 | UP | 23.48 | 131.55 | 111.27 | 0.0601 |
| CAR 14:1 | UP | 20.41 | 79.15 | 62.71 | 0.0642 |
| CAR 16:1 | UP | 17.92 | 33.96 | 27.29 | 0.0672 |
| Hex3Cer 18:1;O2/16:0 | UP | 10.86 | 73.21 | 62.06 | 0.0730 |
| Hex3Cer 18:1;O2/24:1 | UP | 15.24 | 17.70 | 14.35 | 0.0755 |
| CAR 12:1 | UP | 21.47 | 71.70 | 55.62 | 0.0935 |
| CAR 14:2 | UP | 12.96 | 35.18 | 27.11 | 0.1101 |
| CAR 16:1;O | UP | 10.09 | 4.78 | 4.33 | 0.1512 |
| FA 22:5 | UP | 9.69 | 4948.84 | 4242.47 | 0.1535 |
| FA 22:4 | UP | 16.31 | 3810.14 | 3540.19 | 0.1781 |
| CAR 14:0;O | UP | 4.32 | 6.41 | 5.67 | 0.3455 |
| HexCer 18:1;O2/16:0 | UP | 15.62 | 452.28 | 432.98 | 0.3877 |
| SM 18:0;O2/18:0 | UP | 14.18 | 1471.20 | 1263.45 | 0.3877 |
| TG(56:7) [NL-20:4] | UP | -7.87 | 4105.22 | 4056.72 | 0.3925 |
| LPC 20:4 [+OH] | UP | 19.93 | 4.72 | 4.34 | 0.4156 |
| FA 16:1 | UP | 17.64 | 11235.44 | 8865.05 | 0.4234 |
| Cer 18:0;O2/18:0 | UP | 5.66 | 179.08 | 171.18 | 0.4255 |
| Cer 18:1;O2/16:0 | UP | 17.40 | 233.90 | 225.42 | 0.4255 |
| FA 18:1 | UP | 5.13 | 239723.57 | 221455.96 | 0.4450 |
| LPC O-22:1 | UP | -2.29 | 66.49 | 66.48 | 0.4450 |
| PC 36:4 [+OH] | UP | 16.94 | 89.31 | 84.71 | 0.4480 |
| PS 36:2 | UP | 124.69 | 117.52 | 110.97 | 0.4491 |
| TG(54:6) [NL-20:4] | UP | -15.41 | 8161.08 | 8062.44 | 0.4539 |
| TG(58:10) [NL-22:6] | UP | -1.32 | 449.03 | 444.52 | 0.4957 |
| TG(56:8) [NL-22:6] | UP | -5.53 | 2652.54 | 2638.46 | 0.5235 |
| CAR 18:1 | UP | 12.55 | 168.56 | 160.22 | 0.5350 |
| TG(56:7) [NL-22:6] | UP | -6.24 | 4023.84 | 3828.03 | 0.5746 |
| Cer 17:1;O2/18:0 | UP | 4.55 | 9.34 | 8.80 | 0.5758 |
| Hex2NeuAcCer 18:1;O2/16:0 | UP | 6.42 | 42.95 | 41.63 | 0.5835 |
| PE 17:0_20:4 | UP | 3.83 | 138.12 | 131.31 | 0.5835 |
| PE 18:0_22:5 n3 | UP | 0.88 | 402.16 | 388.47 | 0.6295 |
| LPC O-18:1 | UP | 3.47 | 921.57 | 864.13 | 0.6307 |
| PE 18:0_22:5 n6 | UP | -3.77 | 226.81 | 217.56 | 0.6374 |
| TG(52:3) [NL-16:1] | UP | -3.29 | 34943.41 | 34567.24 | 0.6385 |
| HexCer 18:1;O2/24:1 | UP | 11.05 | 560.11 | 536.60 | 0.6475 |
| FA 20:4 | UP | 11.48 | 17193.73 | 16097.98 | 0.6665 |
| DG 18:1_20:4 | UP | -7.40 | 1392.79 | 1338.85 | 0.6947 |
| CAR 12:0 | UP | -1.28 | 46.24 | 45.52 | 0.6958 |
| CAR 14:0 | UP | 2.98 | 24.55 | 21.72 | 0.7004 |
| PC P-16:0/16:0 | UP | 3.18 | 1331.78 | 1303.82 | 0.7187 |
| DG 18:0_20:4 | UP | 4.77 | 314.76 | 288.76 | 0.7302 |
| PC O-40:5 | UP | -0.93 | 820.27 | 793.51 | 0.7302 |
| CAR 24:1 | UP | 3.62 | 2.05 | 2.05 | 0.7612 |
| SM 18:1;O2/17:0 \| SM 17:1;O2/18:0 | UP | -1.43 | 2955.28 | 2926.93 | 0.7612 |
| PE 16:0_18:2 | UP | -10.87 | 2190.27 | 2148.44 | 0.7646 |
| Cer 18:1;O2/20:0 | UP | 2.47 | 132.37 | 131.91 | 0.7669 |
| SPB 18:1;O2 | UP | 84.90 | 33.77 | 32.40 | 0.7680 |
| Cer 20:1;O2/24:1 | UP | 3.09 | 21.07 | 20.37 | 0.7725 |
| TG(54:4) [NL-18:2] | UP | -6.37 | 61649.93 | 59284.25 | 0.7725 |
| PE 16:0_20:4 | UP | 5.99 | 2440.40 | 2323.17 | 0.7932 |
| PE 18:1/18:1 | UP | 4.15 | 661.32 | 589.88 | 0.8139 |
| HexCer 18:1;O2/20:0 | UP | 4.22 | 56.75 | 55.59 | 0.8184 |
| CAR 22:5;O | UP | 1.03 | 0.70 | 0.67 | 0.8275 |
| Hex3Cer 18:1;O2/24:0 | UP | 1.43 | 18.71 | 18.49 | 0.8410 |
| TG(53:2) [NL-17:1] | UP | -14.22 | 583.01 | 564.56 | 0.8410 |
| Cer 18:0;O2/24:1 | UP | -2.23 | 194.08 | 186.20 | 0.8490 |
| PE 18:0_22:4 | UP | 16.79 | 171.62 | 166.84 | 0.8626 |
| TG(54:3) [NL-18:1] | UP | -0.08 | 99721.94 | 96793.51 | 0.8636 |
| Cer 18:1;O/18:0 | UP | -9.07 | 90.77 | 84.47 | 0.8667 |
| Cer 18:1;O/24:1 | UP | -2.34 | 422.49 | 415.62 | 0.8667 |
| FA 16:0 | UP | -1.92 | 72722.91 | 72459.93 | 0.9121 |
| FA 20:5 | UP | 4.35 | 2206.16 | 2123.43 | 0.9248 |
| Hex2NeuAcCer 18:1;O2/24:1 | UP | 5.65 | 23.15 | 22.98 | 0.9287 |
| TG(52:3) [SIM] | UP | -1.58 | 27370.03 | 25749.08 | 0.9287 |
| Cer 18:0;O2/20:0 | UP | 4.15 | 84.57 | 82.96 | 0.9376 |
| PI 18:0_22:5 n6 | UP | 3.61 | 306.36 | 297.79 | 0.9376 |
| CAR 20:3;O | UP | -1.43 | 1.02 | 1.01 | 0.9396 |
| PE 16:0_16:0 | UP | 12.34 | 47.00 | 46.68 | 0.9596 |
| DG 18:1_22:5 | UP | -2.03 | 490.54 | 474.75 | 0.9614 |
| SM 18:1;O2/18:0 \| SM 16:1;O2/20:0 | UP | 1.71 | 21080.70 | 21073.26 | 0.9651 |
| TG(58:8) [NL-22:6] | UP | 5.51 | 99.19 | 98.55 | 0.9908 |
| Hex2NeuAcCer 18:1;O2/18:0 | UP | 3.10 | 14.66 | 14.40 | 0.9926 |
| SM 44:3;O2 | UP | 6.48 | 240.09 | 234.19 | 0.9934 |
| TG(58:9) [NL-22:6] | UP | 10.27 | 728.61 | 657.72 | 0.9966 |
| ^A^ p-values were calculated using Mann Whitney U-Test  ^B^ mean percent different = (mean_case_ – mean_control_ / mean_control_) x 100 | | | | | |

| **Model** | **Continuous NRI (95% CI)** | **IDI (95% CI)** |
| --- | --- | --- |
| Covariates Alone | Ref. | Ref. |
| Covariates + Lipid Score | 1.07 (0.82-1.31) | 0.20 (0.16-0.25) ^a^ |
| ^a^ Confidence interval for IDI was calculated using standard error estimation rather than bootstrapping, due to model convergence issues. | | |

**Supplementary Table 2.** Net reclassification index (NRI) and Integrated Discrimination Improvement (IDI) comparing prediction models using known risk factors for malnutrition alone and combined with Lipid Malnutrition Risk Score among 180 participants with head and neck, gastrointestinal, or lung cancers treated at the Huntsman Cancer Institute (**HCI**).

**Supplementary Table 3.** Complete results from Lipid Ontology (LION) enrichment analysis, including all tested lipid terms and biophysical properties. The analysis was performed using plasma lipidomics data from 180 participants with head and neck, gastrointestinal, or lung cancers treated at the Huntsman Cancer Institute (**HCI**). Participants were categorized by malnutrition risk status using the Malnutrition Screening Tool (MST), with cases defined as an MST score ≥2 and controls as MST score = 0.

| **Description** | **p-value** | **FDR q-value** | **ES** | **Regulated** |
| --- | --- | --- | --- | --- |
| headgroup with positive charge / zwitter-ion | 5.7e-13 | 7.07e-11 | -0.29 | DOWN |
| glycerophosphocholines [GP01] | 6.5e-12 | 4.03e-10 | -0.32 | DOWN |
| endoplasmic reticulum (ER) | 1.2e-11 | 4.96e-10 | -0.28 | DOWN |
| glycerolipids [GL] | 2.3e-11 | 7.13e-10 | 0.35 | UP |
| glycerophospholipids [GP] | 1.3e-10 | 3.22e-09 | -0.26 | DOWN |
| diacylglycerophosphoethanolamines [GP0201] | 1.5e-09 | 3.1e-08 | 0.59 | UP |
| triacylglycerols [GL0301] | 6.3e-09 | 1.12e-07 | 0.33 | UP |
| headgroup with neutral charge | 7.4e-09 | 1.15e-07 | 0.27 | UP |
| monoacylglycerophosphocholines [GP0105] | 1e-08 | 1.38e-07 | -0.44 | DOWN |
| triradylglycerols [GL03] | 1.2e-08 | 1.49e-07 | 0.32 | UP |
| positive intrinsic curvature | 2e-07 | 2.25e-06 | -0.31 | DOWN |
| membrane component | 3.2e-07 | 3.31e-06 | -0.22 | DOWN |
| fatty acids [FA] | 8.4e-07 | 7.44e-06 | 0.81 | UP |
| fatty acids and conjugates [FA01] | 8.4e-07 | 7.44e-06 | 0.81 | UP |
| lysoglycerophospholipids | 1e-06 | 8.27e-06 | -0.34 | DOWN |
| very low bilayer thickness | 2.5e-06 | 1.94e-05 | -0.68 | DOWN |
| polyunsaturated fatty acid | 3.1e-06 | 2.26e-05 | -0.21 | DOWN |
| 1-(1z-alkenyl),2-acylglycerophosphoethanolamines [GP0203] | 6.6e-06 | 4.55e-05 | -0.39 | DOWN |
| diacylglycerophosphocholines [GP0101] | 7.5e-06 | 4.89e-05 | -0.32 | DOWN |
| lipid storage | 9.4e-06 | 5.55e-05 | 0.24 | UP |
| lipid droplet | 9.4e-06 | 5.55e-05 | 0.24 | UP |
| neutral intrinsic curvature | 1.6e-05 | 8.63e-05 | -0.24 | DOWN |
| contains vinyl ether bond (plasmalogen) | 1.6e-05 | 8.63e-05 | -0.31 | DOWN |
| 1-alkyl,2-acylglycerophosphoethanolamines [GP0202] | 3.8e-05 | 0.000196 | -0.59 | DOWN |
| fatty acid with 3 double bonds | 4.1e-05 | 0.000203 | -0.4 | DOWN |
| diacylglycerophosphoserines [GP0301] | 7.9e-05 | 0.000377 | 0.92 | UP |
| diacylglycerols [GL0201] | 8.8e-05 | 0.000404 | 0.49 | UP |
| above average lateral diffusion | 0.00014 | 0.00062 | -0.37 | DOWN |
| very high lateral diffusion | 0.00015 | 0.000641 | -0.56 | DOWN |
| d18:1 (sphingosine) | 0.00021 | 0.000868 | -0.67 | DOWN |
| C20:3 | 0.0003 | 0.0012 | -0.45 | DOWN |
| below average bilayer thickness | 0.00031 | 0.0012 | -0.37 | DOWN |
| fatty acid with more than 3 double bonds | 0.00109 | 0.00405 | -0.17 | DOWN |
| C18:2 | 0.00111 | 0.00405 | -0.25 | DOWN |
| fatty acid with 2 double bonds | 0.00121 | 0.00429 | -0.24 | DOWN |
| fatty acid with more than 18 carbons | 0.00141 | 0.00486 | -0.16 | DOWN |
| fatty acid with 19-21 carbons | 0.00172 | 0.00576 | -0.19 | DOWN |
| fatty acid with 20 carbons | 0.00235 | 0.00767 | -0.19 | DOWN |
| C20:5 | 0.00312 | 0.00977 | -0.46 | DOWN |
| C18:1 | 0.00315 | 0.00977 | 0.19 | UP |
| saturated fatty acid | 0.00355 | 0.0107 | -0.15 | DOWN |
| 1z-alkenylglycerophosphocholines [GP0107] | 0.00551 | 0.0163 | -0.65 | DOWN |
| dihexosylceramides | 0.00593 | 0.0171 | 0.54 | UP |
| monounsaturated fatty acid | 0.00692 | 0.0195 | 0.15 | UP |
| high bilayer thickness | 0.00981 | 0.027 | 0.41 | UP |
| below average lateral diffusion | 0.01021 | 0.0275 | 0.41 | UP |
| C22:6 | 0.01143 | 0.0302 | -0.26 | DOWN |
| headgroup with negative charge | 0.01215 | 0.0314 | 0.27 | UP |
| C15:0 | 0.01285 | 0.0325 | -0.44 | DOWN |
| monoalkylglycerophosphocholines [GP0106] | 0.01405 | 0.0348 | 0.48 | UP |
| simple glc series [SP0501] | 0.01451 | 0.0353 | 0.34 | UP |
| low lateral diffusion | 0.01603 | 0.0382 | 0.4 | UP |
| very low transition temperature | 0.01985 | 0.0464 | -0.39 | DOWN |
| fatty acid with 6 double bonds | 0.02034 | 0.0467 | -0.24 | DOWN |
| fatty acid with 18 carbons or less | 0.0215 | 0.0485 | -0.12 | DOWN |
| below average transition temperature | 0.02493 | 0.0552 | -0.24 | DOWN |
| above average transition temperature | 0.02593 | 0.0564 | 0.5 | UP |
| negative intrinsic curvature | 0.02641 | 0.0565 | 0.13 | UP |
| fatty acid with 3-5 double bonds | 0.02998 | 0.063 | -0.14 | DOWN |
| diacylglycerophosphoinositols [GP0601] | 0.03235 | 0.0669 | -0.33 | DOWN |
| glycerophosphoinositols [GP06] | 0.03466 | 0.0705 | -0.3 | DOWN |
| contains ether-bond | 0.03626 | 0.0725 | -0.25 | DOWN |
| C22:4 | 0.04029 | 0.0793 | 0.39 | UP |
| C18:3 | 0.04295 | 0.0832 | -0.39 | DOWN |
| steryl esters [ST0102] | 0.04519 | 0.0853 | -0.26 | DOWN |
| monoacylglycerophosphoethanolamines [GP0205] | 0.04541 | 0.0853 | -0.38 | DOWN |
| fatty acid with 13-15 carbons | 0.04844 | 0.0897 | -0.27 | DOWN |
| plasma membrane | 0.05092 | 0.0929 | -0.14 | DOWN |
| average lateral diffusion | 0.05412 | 0.0973 | 0.28 | UP |
| high transition temperature | 0.05774 | 0.102 | 0.48 | UP |
| endosome/lysosome | 0.06522 | 0.112 | -0.16 | DOWN |
| C24:1 | 0.06524 | 0.112 | 0.68 | UP |
| lipid-mediated signalling | 0.06619 | 0.112 | -0.12 | DOWN |
| ceramide phosphocholines (sphingomyelins) [SP0301] | 0.06952 | 0.116 | -0.2 | DOWN |
| golgi apparatus | 0.07749 | 0.127 | -0.19 | DOWN |
| DG(36:4) | 0.07785 | 0.127 | 0.66 | UP |
| average bilayer thickness | 0.08621 | 0.139 | 0.28 | UP |
| C19:0 | 0.11976 | 0.19 | -0.62 | DOWN |
| fatty acid with 17 carbons | 0.12836 | 0.201 | -0.2 | DOWN |
| C14:0 | 0.13315 | 0.206 | -0.34 | DOWN |
| high lateral diffusion | 0.1409 | 0.213 | -0.24 | DOWN |
| C22:0 | 0.1411 | 0.213 | -0.53 | DOWN |
| N-acylsphingosines (ceramides) [SP0201] | 0.15697 | 0.233 | 0.16 | UP |
| C20:4 | 0.15752 | 0.233 | -0.16 | DOWN |
| fatty acid with 22 carbons | 0.16692 | 0.242 | -0.13 | DOWN |
| sphingolipids [SP] | 0.16766 | 0.242 | -0.11 | DOWN |
| 1z-alkenylglycerophosphoethanolamines [GP0207] | 0.16992 | 0.242 | 0.51 | UP |
| C24:0 | 0.17316 | 0.244 | -0.46 | DOWN |
| C20:1 | 0.17851 | 0.249 | 0.33 | UP |
| fatty acid with 16-18 carbons | 0.18278 | 0.252 | -0.08 | DOWN |
| C17:1 | 0.19191 | 0.261 | -0.27 | DOWN |
| 1-(1z-alkenyl),2-acylglycerophosphocholines [GP0103] | 0.19328 | 0.261 | -0.22 | DOWN |
| d18:2 | 0.21359 | 0.285 | -0.34 | DOWN |
| fatty acid with 24 carbons | 0.21655 | 0.286 | 0.29 | UP |
| C22:1 | 0.22361 | 0.292 | 0.55 | UP |
| fatty acid with 5 double bonds | 0.24466 | 0.316 | -0.17 | DOWN |
| C20:2 | 0.24921 | 0.319 | -0.47 | DOWN |
| PE(P-38:4) | 0.2659 | 0.336 | -0.53 | DOWN |
| fatty acid with 14 carbons | 0.27717 | 0.347 | -0.27 | DOWN |
| fatty acid with 18 carbons | 0.30536 | 0.379 | -0.08 | DOWN |
| C26:0 | 0.39943 | 0.486 | -0.47 | DOWN |
| fatty acid with 22-24 carbons | 0.4035 | 0.486 | -0.1 | DOWN |
| low bilayer thickness | 0.40362 | 0.486 | -0.2 | DOWN |
| hexosylceramides | 0.41364 | 0.493 | 0.24 | UP |
| C17:0 | 0.42319 | 0.5 | -0.2 | DOWN |
| fatty acid with less than 2 double bonds | 0.43542 | 0.509 | -0.07 | DOWN |
| d18:0 (dihydrosphingosine) | 0.45179 | 0.524 | -0.3 | DOWN |
| fatty acid with 4 double bonds | 0.45922 | 0.527 | -0.11 | DOWN |
| 1-alkyl,2-acylglycerophosphocholines [GP0102] | 0.48638 | 0.553 | 0.19 | UP |
| DG(38:6) | 0.49455 | 0.557 | 0.44 | UP |
| C18:0 | 0.52716 | 0.589 | -0.1 | DOWN |
| low transition temperature | 0.53805 | 0.596 | -0.16 | DOWN |
| average transition temperature | 0.56824 | 0.624 | 0.17 | UP |
| glycerophosphoethanolamines [GP02] | 0.59075 | 0.64 | -0.08 | DOWN |
| C19:1 | 0.59524 | 0.64 | 0.26 | UP |
| mitochondrion | 0.59826 | 0.64 | -0.08 | DOWN |
| C20:0 | 0.70137 | 0.743 | -0.19 | DOWN |
| C16:0 | 0.73937 | 0.777 | 0.09 | UP |
| fatty acid with 26 carbons | 0.75227 | 0.784 | -0.31 | DOWN |
| monoacylglycerophosphoinositols [GP0605] | 0.77086 | 0.79 | -0.34 | DOWN |
| C22:5 | 0.771 | 0.79 | 0.14 | UP |
| fatty acid with 16 carbons | 0.92172 | 0.93 | 0.06 | UP |
| C16:1 | 0.92287 | 0.93 | 0.1 | UP |
| fatty acids with 19 carbons | 0.95581 | 0.956 | 0.14 | UP |

**Supplementary Table 4.** Elastic net-derived weights and univariate associations with malnutrition risk (Malnutrition Screening Tool score = 0 versus ≥2) for the lipids consistently selected (>90% of iterations) by Elastic net regression offset for potential confounders ^A^ and GDF15 levels among 180 participants with head and neck, gastrointestinal, or lung cancers treated at the Huntsman Cancer Institute (HCI).

| **Lipid** | **Main Lipid Class** | **ß Coefficient** | **OR (95% CI)** |
| --- | --- | --- | --- |
| CE 20:0 | Sterol Lipids | -0.08 | 0.47 (0.31-0.71) |
| Hex2Cer 18:1;O2/20:0 | Sphingolipids | 0.27 | 1.49 (1.09-2.03) |
| LPC 26:0/0:0 | Glycerophospholipids | -0.24 | 0.42 (0.28-0.64) |
| LPI 18:2/0:0 | Glycerophospholipids | -0.19 | 0.60 (0.42-0.85) |
| PC 34:5 | Glycerophospholipids | -0.07 | 0.38 (0.25-0.59) |
| PC 40:8 | Glycerophospholipids | -0.05 | 0.36 (0.23-0.57) |
| PC P-18:0/18:2 | Glycerophospholipids | -0.09 | 0.48 (0.33-0.69) |
| PE P-18:1/18:2 b | Glycerophospholipids | -0.05 | 0.44 (0.30-0.65) |
| SHexCer 18:1;O2/16:0 | Sphingolipids | -0.01 | 0.41 (0.26-0.63) |
| SM 18:2;O2/23:0 | Sphingolipids | -0.07 | 0.32 (0.20-0.51) |
| TG(O-50:1) [NL-18:1] | Glycerolipids | -0.08 | 0.51 (0.36-0.73) |
| TG(O-50:1) [SIM] | Glycerolipids | -0.07 | 0.59 (0.43-0.82) |
| TG(O-54:3) | Glycerolipids | -0.02 | 0.63 (0.45-0.87) |
| Cholesterol Ester (**CE**); Dihexosylceramide (**Hex2Cer**); Lysophosphatidylcholine (**LPC**); Lysophosphatidylinositol (**LPI**); Phosphatidylcholine (**PC**); Phosphatidylethanolamine (**PE**); Sulfated hexosylceramide (**SHexCer**); Sphingomyelin (**SM**); Triglyceride (**TG**) | | | |

**Supplementary Table 5.** Elastic net-derived weights and univariate associations with malnutrition risk (Malnutrition Screening Tool score = 0 versus ≥2) for the lipids consistently selected (>90% of iterations) by Elastic net regression offset for potential confounders ^A^ and imperfectly matched matching factors (number of freeze/thaws and anticoagulation treatment) among 180 participants with head and neck, gastrointestinal, or lung cancers treated at the Huntsman Cancer Institute (HCI).

| **Lipid** | **Main Lipid Class** | **ß Coefficient** | **OR (95% CI)** |
| --- | --- | --- | --- |
| CE 20:0 | Sterol Lipids | -0.03 | 0.47 (0.31-0.71) |
| Cer 18:2;O2/26:0 | Sphingolipids | -0.11 | 0.44 (0.30-0.65) |
| Hex2Cer 18:1;O2/20:0 | Sphingolipids | 0.33 | 1.49 (1.09-2.03) |
| LPC 26:0/0:0 | Glycerophospholipids | -0.30 | 0.42 (0.28-0.64) |
| LPI 18:2/0:0 | Glycerophospholipids | -0.10 | 0.60 (0.42-0.85) |
| PC 34:5 | Glycerophospholipids | -0.17 | 0.38 (0.25-0.59) |
| PC 40:8 | Glycerophospholipids | -0.13 | 0.36 (0.23-0.57) |
| PE P-18:1/18:2 b | Glycerophospholipids | -0.11 | 0.44 (0.30-0.65) |
| PE P-18.1/20.4 b | Glycerophospholipids | -0.11 | 0.38 (0.24-0.60) |
| PI 16:0/16:0 | Glycerophospholipids | -0.02 | 0.54 (0.36-0.80) |
| SM 18:2;O2/23:0 | Sphingolipids | -0.11 | 0.32 (0.20-0.51) |
| TG(O-50:1) | Glycerolipids | -0.16 | 0.51 (0.36-0.73) |
| Cholesterol Ester (**CE**); Ceramide (**Cer**); Dihexosylceramide (**Hex2Cer**); Lysophosphatidylcholine (**LPC**); Lysophosphatidylinositol (**LPI**); Phosphatidylcholine (**PC**); Phosphatidylethanolamine (**PE**); Phosphatidylinositol (**PI**); Sphingomyelin (**SM**); Triglyceride (**TG**) | | | |

**Supplementary Table 6.** Elastic net-derived weights and univariate associations with malnutrition risk (Malnutrition Screening Tool (MST) score = 0 versus ≥2) for the lipids consistently selected (>90% of iterations) by Elastic net regression offset for potential confounders ^A^ among participants with head and neck, gastrointestinal, or lung cancers treated at the Huntsman Cancer Institute (HCI), excluding participants with a cancer diagnosed before 2021 (the year of MST implementation) (n = 152).

| **Lipid** | **Main Lipid Class** | **ß Coefficient** | **OR (95% CI)** |
| --- | --- | --- | --- |
| CE 20:4 | Sterol lipids | -0.17 | 0.34 (0.20-0.58) |
| Cer 18:2;O2/23:0 | Sphingolipids | -0.15 | 0.41 (0.27-0.63) |
| Cer 18:2;O2/24:0 | Sphingolipids | -0.29 | 0.45 (0.31-0.68) |
| Cer 18:2;O2/26:0 | Sphingolipids | -0.13 | 0.47 (0.31-0.70) |
| Hex2Cer 18:1;O2/20:0 | Sphingolipids | 0.47 | 1.50 (1.06- 2.12) |
| LPC 26:0/0:0 | Glycerophospholipids | -0.58 | 0.32 (0.19-0.55) |
| LPI 18:2/0:0 | Glycerophospholipids | -0.22 | 0.54 (0.36-0.82) |
| PC P-16:0/18:2 | Glycerophospholipids | -0.09 | 0.49 (0.34-0.73) |
| PC P-18:0/18:2 | Glycerophospholipids | -0.38 | 0.49 (0.33-0.72) |
| PI 16:0/16:0 | Glycerophospholipids | -0.27 | 0.51 (0.33-0.80) |
| SM 18:2;O2/23:0 | Sphingolipids | 0.09 | 0.32 (0.19-0.53) |
| Cholesterol Ester (**CE**); Ceramide (**Cer**); Dihexosylceramide (**Hex2Cer**); Lysophosphatidylcholine (**LPC**); Lysophosphatidylinositol (**LPI**); Phosphatidylcholine (**PC**); Sphingomyelin (**SM**); Triglyceride (**TG**); Phosphatidylinositol (**PI**) | | | |
