## Supplementary figures and images for "Identification of circulating lipidomic biomarkers of malnutrition risk among oncology patients in the Total Cancer Care (TCC) Study: a cross-sectional analysis"

### Supplemental Figure 1

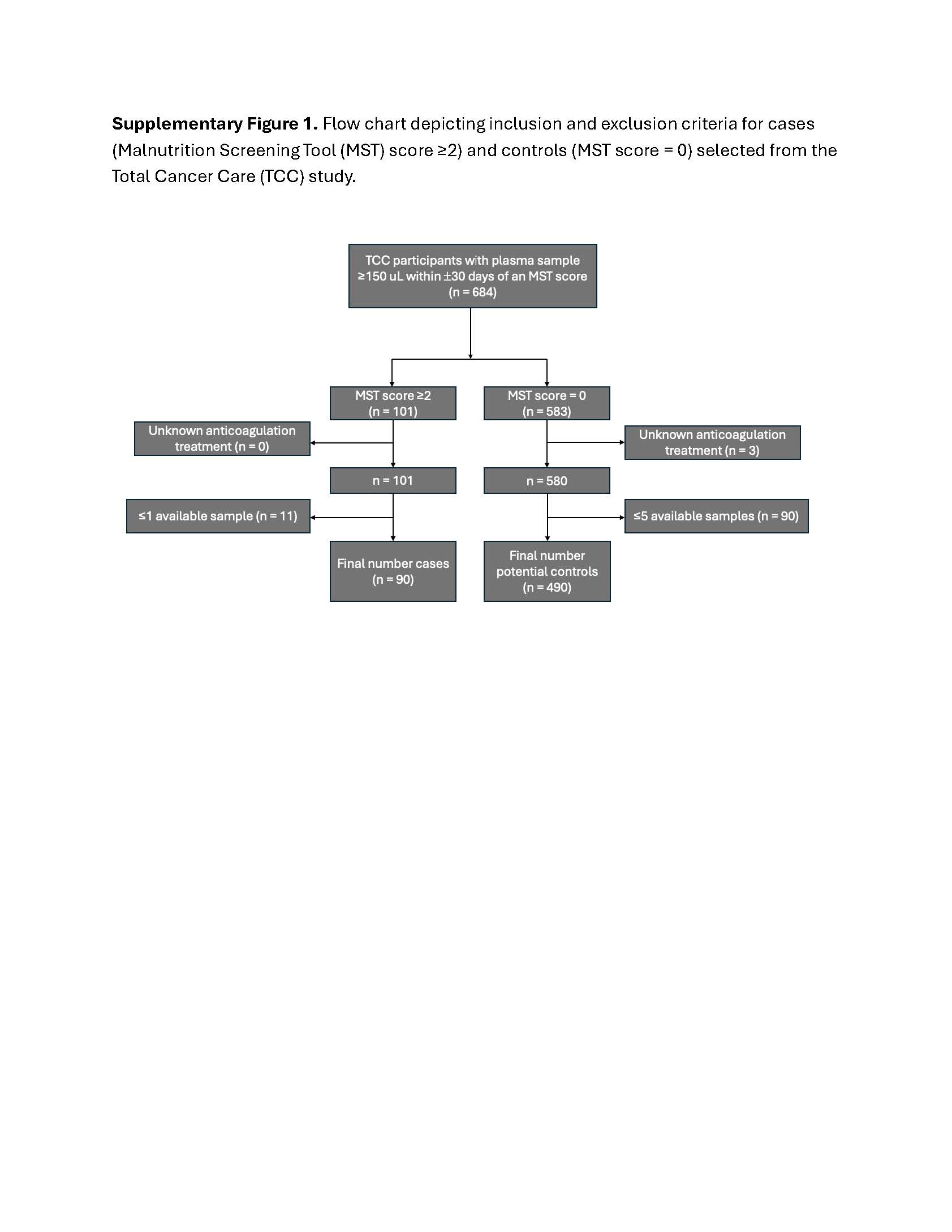
